# Impact of non-pharmaceutical interventions on SARS-CoV-2 outbreaks in English care homes: a modelling study

**DOI:** 10.1101/2021.05.17.21257315

**Authors:** Alicia Roselló, Rosanna C. Barnard, David R. M. Smith, Stephanie Evans, Fiona Grimm, Nicholas G. Davies, Centre for Mathematical Modelling of Infectious Diseases COVID-19 modelling working group, Sarah R. Deeny, Gwenan M. Knight, W. John Edmunds

**Author notes:** Working group members are listed separately. Members of the Centre for Mathematical Modelling of Infectious Diseases COVID-19 modelling working group (random order): Carl A B Pearson, Thibaut Jombart, Simon R Procter, Yung-Wai Desmond Chan, Graham Medley, Joel Hellewell, Billy J Quilty, Yang Liu, Stefan Flasche, Sam Abbott, Rosalind M Eggo, Rachel Lowe, Yalda Jafari, Christopher I Jarvis, Naomi R Waterlow, James D Munday, Amy Gimma, Nikos I Bosse, Petra Klepac, Akira Endo, Adam J Kucharski, Rosanna C Barnard, C Julian Villabona-Arenas, Sebastian Funk, Kevin van Zandvoort, W John Edmunds, William Waites, Emily S Nightingale, Sophie R Meakin, Frank G Sandmann, Fabienne Krauer, Nicholas G. Davies, Alicia Showering, Samuel Clifford, Katherine E. Atkins, Matthew Quaife, Kaja AbbasTimothy W Russell, Fiona Yueqian Sun, Mark Jit, Mihaly Koltai, Kiesha Prem, Jiayao Lei, Anna M Foss, Jack Williams, Kathleen O’Reilly, Damien C Tully, Hamish P Gibbs, Oliver Brady. CMMID COVID-19 modelling working group funding statements: CABP (B&MGF: NTD Modelling Consortium OPP1184344, FCDO/Wellcome Trust: Epidemic Preparedness Coronavirus research programme 221303/Z/20/Z), TJ (Global Challenges Research Fund: ES/P010873/1, UK Public Health Rapid Support Team, NIHR: Health Protection Research Unit for Modelling Methodology HPRU-2012-10096, UK MRC: MC_PC_19065), SRP(B&MGF: INV-016832), GFM (B&MGF: NTD Modelling Consortium OPP1184344), JH (Wellcome Trust: 210758/Z/18/Z), BJQ (NIHR: 16/137/109, NIHR: 16/136/46, B&MGF: OPP1139859), YL (B&MGF: INV-003174, NIHR: 16/137/109, European Commission: 101003688, UK MRC: MC_PC_19065), SFlasche (Wellcome Trust: 208812/Z/17/Z), SA (Wellcome Trust: 210758/Z/18/Z), RME (HDR UK: MR/S003975/1, UK MRC: MC_PC_19065, NIHR: NIHR200908), RL (Royal Society: Dorothy Hodgkin Fellowship), YJ UKRI: MR/V028456/1, CIJ (Global Challenges Research Fund: ES/P010873/1), NRW(MRC: MR/N013638/1), JDM (Wellcome Trust: 210758/Z/18/Z), AG (European Commission: 101003688), NIB (HPRU: NIHR200908), PK (Royal Society: RP\EA\180004, European Commission: 101003688), AE (Nakajima Foundation), AJK (Wellcome Trust: 206250/Z/17/Z, NIHR: NIHR200908), RCB (European Commission: 101003688), SFunk (Wellcome Trust: 210758/Z/18/Z), KvZ (Elrha R2HC/UK FCDO/Wellcome Trust/NIHR, FCDO/Wellcome Trust: Epidemic Preparedness Coronavirus research programme 221303/Z/20/Z), WJE (European Commission: 101003688, UK MRC: MC_PC_19065, NIHR: PR-OD-1017-20002), WW (MRC: MR/V027956/1), ESN (B&MGF: OPP1183986), SRM (Wellcome Trust: 210758/Z/18/Z), FGS (NIHR: NIHR200929), FK (Innovation Fund: 01VSF18015, Wellcome Trust: UNS110424), NGD (UKRI Research England, NIHR: NIHR200929, UK MRC: MC_PC_19065), SC (Wellcome Trust: 208812/Z/17/Z, UK MRC: MC_PC_19065), MQ (ERC Starting Grant: #757699, B&MGF: INV-001754), KA (BMGF: INV-016832; OPP1157270), TWR (Wellcome Trust: 206250/Z/17/Z), FYS (NIHR: 16/137/109), MJ (B&MGF: INV-003174, NIHR: 16/137/109, NIHR: NIHR200929, European Commission: 101003688), MK (Wellcome Trust 221303/Z/20/Z), KP (B&MGF: INV-003174, European Commission: 101003688), JYL Gates (INV-003174), KO’R (B&MGF: OPP1191821), HPG (EDCTP2: RIA2020EF-2983-CSIGN, UK DHSC/UK Aid/NIHR: PR-OD-1017-20001), OJB (Wellcome Trust: 206471/Z/17/Z).

## Abstract

**Background:** COVID-19 outbreaks are still occurring in English care homes despite the non-pharmaceutical interventions (NPIs) in place.

**Methods:** We developed a stochastic compartmental model to simulate the spread of SARS-CoV-2 within an English care home. We quantified the outbreak risk under the NPIs already in place, the role of community prevalence in driving outbreaks, and the relative contribution of all importation routes into the care home. We also considered the potential impact of additional control measures, namely: increasing staff and resident testing frequency, using lateral flow antigen testing (LFD) tests instead of PCR, enhancing infection prevention and control (IPC), increasing the proportion of residents isolated, shortening the delay to isolation, improving the effectiveness of isolation, restricting visitors and limiting staff to working in one care home.

**Findings:** The model suggests that importation of SARS-CoV-2 by staff, from the community, is the main driver of outbreaks, that importation by visitors or from hospitals is rare, and that the past testing strategy (monthly testing of residents and daily testing of staff by PCR) likely provides negligible benefit in preventing outbreaks. Daily staff testing by LFD was 39% (95% 18-55%) effective in preventing outbreaks at 30 days compared to no testing.

**Interpretation:** Increasing the frequency of testing in staff and enhancing IPC are important to preventing importations to the care home. Further work is needed to understand the impact of vaccination in this population, which is likely to be very effective in preventing outbreaks.

**Funding:** The National Institute for Health Research, European Union Horizon 2020, Canadian Institutes of Health Research, French National Research Agency, UK Medical Research Council. The World Health Organisation funded the development of the COS-LTCF Shiny application.

**Research in Context:** *Evidence before this study:* Care homes have been identified as being at increased risk of COVID-19 outbreaks, and a number of modelling studies have considered the transmission dynamics of SARS-CoV-2 in this setting. We searched the PubMed database and bioRxiv and medRxiv’s COVID-19 SARS-CoV-2 preprints for English-language articles on the 11th May 2021, with the search terms (“COVID-19” OR “SARS-CoV-2” OR “coronavirus”) AND (“care home” OR “LTCF” OR “long term care facility” OR “nursing home”) AND (“model”). In addition to these searches, we identified articles relevant to this work through informal networks. These searches returned 87 studies, of which 12 explicitly modelled SARS-CoV-2 transmission within care homes and explored the effectiveness of non-pharmaceutical interventions in these settings. These studies employed a number of modelling approaches (agent-based and compartmental models) and considered various strategies for mitigating epidemic spread within care homes. Only one of these studies modelled care homes in England, but didn’t consider individual care homes as separate entities (transmission between residents in separate facilities was equally likely as within one facility) and only modelled one intervention within the care home: the effect of restricting visitors. Another study modelled a different type of long-term care facility, a rehabilitation facility in France. Other studies modelled care homes in Canada, Scotland, and the US. These modelled care homes were larger than the average English care home. Only one study included importation of SARS-CoV-2 to care homes from hospitals through resident hospitalisation.

*Added value of this study:* We developed a stochastic compartmental model describing the transmission dynamics of SARS-CoV-2 within English care homes. This study is the first to assess the relative importance of all SARS-CoV-2 importation routes to care homes (including resident hospitalisation) and to quantify the impact of a range of non-pharmaceutical interventions against SARS-CoV-2 particularly for English care homes. We found that community prevalence, through staff importations, was the main driver of outbreaks in care homes at 30 days, not importation from hospital visits nor by visitors. In line with this, we found daily testing of staff to be the most effective testing strategy in preventing outbreaks. We show the previous testing strategy (PCR testing residents once every 28 days and staff once a week) to be ineffective in preventing outbreaks and suggest that more frequent testing of staff is required. Restricting visitors bore little effect on the probability of an outbreak occurring by day 30. Interventions focusing on decreasing the transmission of SARS-CoV-2 in the care home were the most effective in reducing the frequency of outbreaks. We provide a Shiny application for users to explore alternative care home characteristics, outbreak characteristics and interventions.

*Implications of all the available evidence:* Preventing the importation of SARS-CoV-2 to care homes from the community through staff is key to preventing outbreaks. Infection prevention and control (IPC) measures targeting transmission within the care home and frequent testing of staff, ideally daily, are the most effective strategies considered. Many care homes in England are currently unable to meet the additional workload daily testing would entail, therefore additional support should be considered to enable these measures. Allowing visitors should be considered given their general positive contribution to residents’ physical and mental health and likely negligible contribution to outbreaks.

## Introduction

Care homes have borne a large burden of the COVID-19 pandemic. A study pooling data from 26 countries found that 41% of all COVID-19 deaths were in care home residents.^1^ In England there were an estimated 30,500 excess death registrations in care home residents between the 23rd March and the 19th of June 2020, compared to the same time period during previous years 2017-2019.^2^ During the third week of January 2021, 45% of COVID-19 deaths in England and Wales were in care home residents, suggesting the measures in place in care homes are not sufficient to prevent and suppress outbreaks.^3^

Approximately 460,000 residents are registered as living in around 15,000 care homes in England, of which around 6,000 provide care exclusively for older people (around 230,000 beds)^4^. This analysis focuses on the latter. Within this manuscript we distinguish between nursing care homes, which provide 24h nursing care (approximately 35% of care homes that provide care to older people in England^4^), and residential care homes, which do not. In practice, this distinction may not be as clear as some nursing care homes may only provide nursing care to some of their residents.

Many factors put care homes at increased risk of infectious disease outbreaks. Firstly, implementing infection prevention and control (IPC) measures in these settings is difficult, since these facilities are residents’ homes and as such frequently contain soft furnishings and shared living spaces. Many residents have high levels of personal care needs due to their clinical conditions which require close and frequent contact with staff, making it impossible for residents and staff to adhere to IPC and physical distancing measures. Isolation capability is also limited for residents with ambulatory dementia. During the COVID-19 pandemic in the UK, IPC has been hindered further by varied access to personal protective equipment (PPE) and testing, as well as a lack of staff sick pay.^5^ Secondly, in England care homes are closely linked to hospitals, with on average 1 hospital admission per resident per year in 2016/17,^6^ making this setting vulnerable to importations from hospital. Thirdly, care homes are closely linked to the community through care home staff and through visitors. Staff working across several care homes could also enhance the spread of SARS-CoV-2 between care homes.^5^

The high burden of COVID-19 mortality seen in care home settings emphasises the need for studies to explore the drivers of SARS-CoV-2 outbreaks as well as to identify effective mitigation and control measures. National COVID-19 guidelines for care homes in England are outlined in Supplementary material A0. In this study we use a mathematical model to simulate spread of SARS-CoV-2 in English care homes with baseline interventions in place. We aim to quantify care home outbreak risk in terms of (i) the baseline scenario, (ii) community prevalence, (iii) the relative contribution of different importation routes, and (iv) the potential reductions provided by additional non-pharmaceutical interventions.

## Methods

### Model overview

We used a stochastic compartmental model to simulate the transmission of SARS-CoV-2 among residents and staff in English care homes (see Supplementary material A1 for details). Two types of facility were considered: a residential care home with 29 beds and 29 members of staff and a nursing care home with 47 beds and 94 members of staff. These resident numbers represent the median bed numbers in these facilities in England^4^ and assumed staff to resident ratios (see Figure S1).^7,8^ We included resident hospitalisation (see schematic in Figure 1), testing of residents and staff, isolation of residents, absence and replacement of staff (see Supplementary material A1), and resident death. The pathways by which residents and staff may become infected are shown in Figure S2. The three routes of SARS-CoV-2 importation into the care home are: from the community or another care home through staff, from the community through visitors, and from hospital through residents. With the exception of the frequency of testing, which was fixed, parameter values were drawn randomly from their respective distributions (see Table S1). Mortality and hospitalisation dynamics are described in detail in Supplementary material A2.

**Figure 1.**
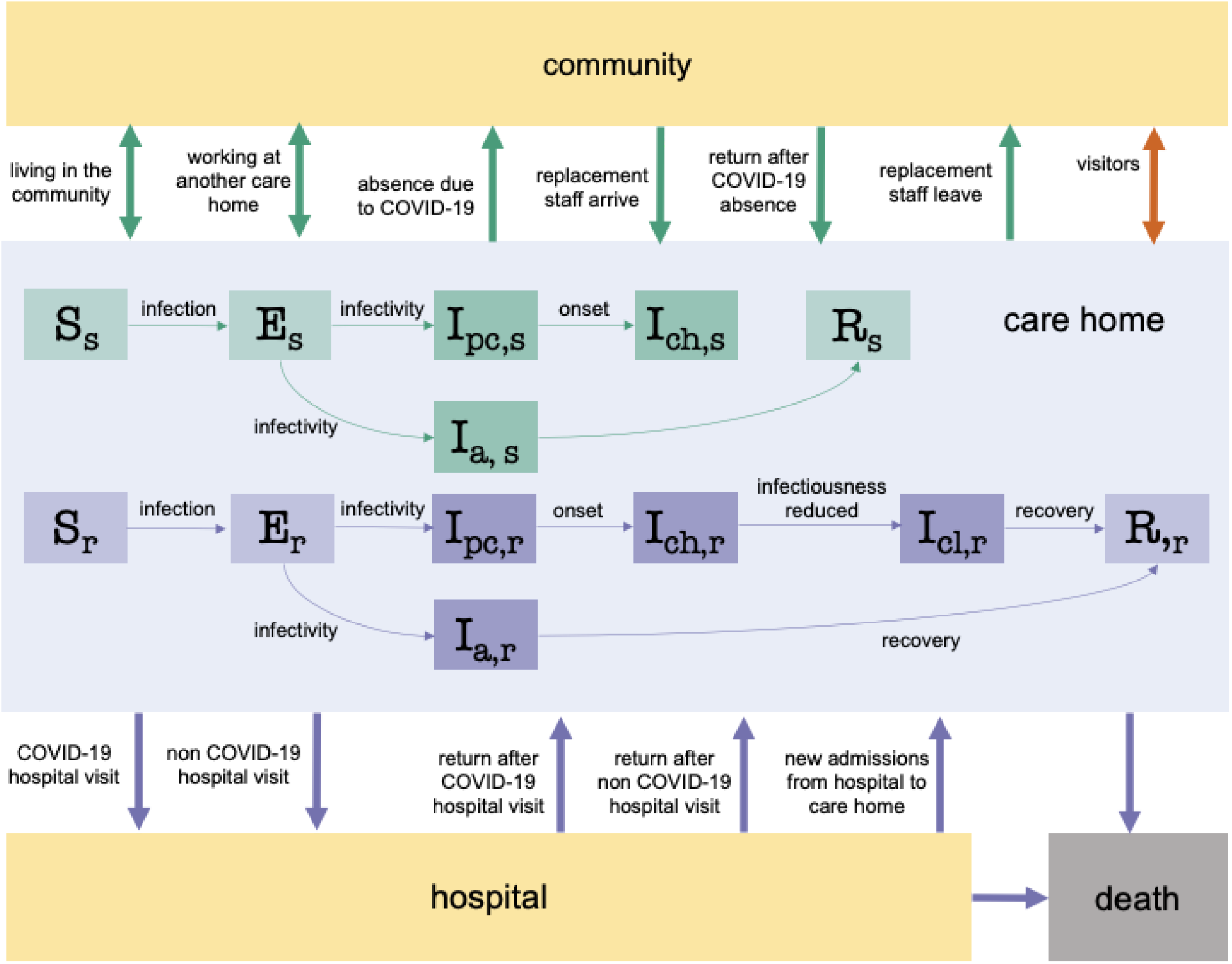
Model schematic of the SARS-CoV-2 infection and disease process in residents and staff. Residents were classified into susceptible (S_r_), exposed (E_r_), infectious asymptomatic (I_a,r_), infectious preclinical (I_pc,r_), infectious clinical with high infectiousness (I_ch,r_), infectious clinical with low infectiousness (I_cl,r_), and recovered (R_r_) compartments. Staff were classified into susceptible (S_s_), exposed (E_s_), infectious asymptomatic (I_a,s_), infectious preclinical (I_pc,s_), infectious clinical with high infectiousness (I_ch,s_), and recovered (R_s_) compartments. Darker shades denote compartments that contribute towards the force of infection. Resident movements are denoted by bold purple arrows, staff movements are denoted by bold green arrows and visiting is denoted by a bold orange arrow. Residents exit the care home due to hospital visits for COVID-19 and non-COVID-19 reasons or as a result of death. Residents enter the care home from the hospital following a COVID-19 admission, a non-COVID-19 admission or as a new admission. Within-hospital transmission dynamics were not modelled explicitly. Flows of new care home residents arriving from the community and care home residents moving into the community are assumed to be negligible during the pandemic and thus are not considered in the model. Staff are assumed to live in the community, and a small proportion of staff work at another care home. Staff may become absent because of COVID-19 symptoms or a positive test, and return to the care home recovered. Absent staff may be replaced by a secondary pool of staff, who in turn leave the care home as the original staff return from their absence.

### Testing and isolation

In the baseline scenario we assumed that, upon presentation of COVID-19 symptoms, a mean of 90% (95% 81-97%) of residents were tested and (if positive and isolation was possible) were isolated within a day of symptom onset. Not all symptomatic residents were tested due to the inability to swab some residents (e.g. agitated residents with severe dementia). On average, we assumed 85% (95% 76-93%) of residents without symptoms were tested every 28 days and 95% (95% 85-97%) of staff without symptoms were tested every 7 days (reflecting the previous testing policy in England^9^). In addition, all residents who were hospitalised were tested in hospital before returning to the care home.

In the baseline scenario, we assumed all tests carried out were using laboratory RT-PCR. PCR testing was assumed to have 100% specificity and, a mean of 90% (95% 88-92%) sensitivity when carried out in hospital^10,11^ (ie. it detected on average 90% of infectious individuals). There is no PCR sensitivity data specific to the care home setting. We assumed sensitivity dropped to 80% (95% 72-88%) when carried out in the care home (as healthcare workers are better trained to carry out these tests than care home staff). The mean delay to isolation/absence (if it occurred) in residents/staff without symptoms tested by PCR was assumed to be 2 days (95% 0.7-3.9). This was defined as the delay between entering an infectious state and isolation/absence, therefore comprising the delay to testing and obtaining testing results. The mean delay to isolation/absence in residents with symptoms was assumed to be 1 day (95% 0.3-1.9). Residents tested in hospital and found positive were assumed to be immediately isolated upon their return to the care home if their hospitalisation was for COVID-19, and after a mean of 2 days (95% 0.7-3.9) if they had been hospitalised for a different reason. LFD tests were explored in alternative testing scenarios and were assumed to have a mean of 70% sensitivity (95% 61-78%), and a mean delay to isolation of 0.25 days (95% 0.08-0.48, given a delay in relocation logistics).^12^

Of those residents testing positive or developing symptoms, on average 80% (95% 66-92%) were isolated (*p*_*i*_). This assumption reflects limited isolation capacity in many care homes.^13^ In the baseline scenario, we assumed that isolation reduced the transmission rate from isolated individuals by an average 75% (47-96%), in addition to the abovementioned reduction across all individuals in care homes with detected outbreaks. Care home resident testing and isolation pathways are further described in Figure S5.

### Scenarios, outputs, sensitivity analysis

All scenarios compared nursing and residential care homes. Three community prevalence scenarios were considered: low (mid-July 2020), medium (baseline scenario, late September 2020), and high (early April 2020). The prevalence scenarios infomed the probability of visitors being infectious, the force of infection to staff from the community, the proportion of staff and replacement staff starting the simulation in each infectious state, the probability of residents being infected in hospital, the non-COVID hospitalisation rate, and the probability of another care home experiencing an outbreak (see Supplementary material A3 for details). Key assumptions of the baseline scenario are listed in Table 1.

**Table 1.**
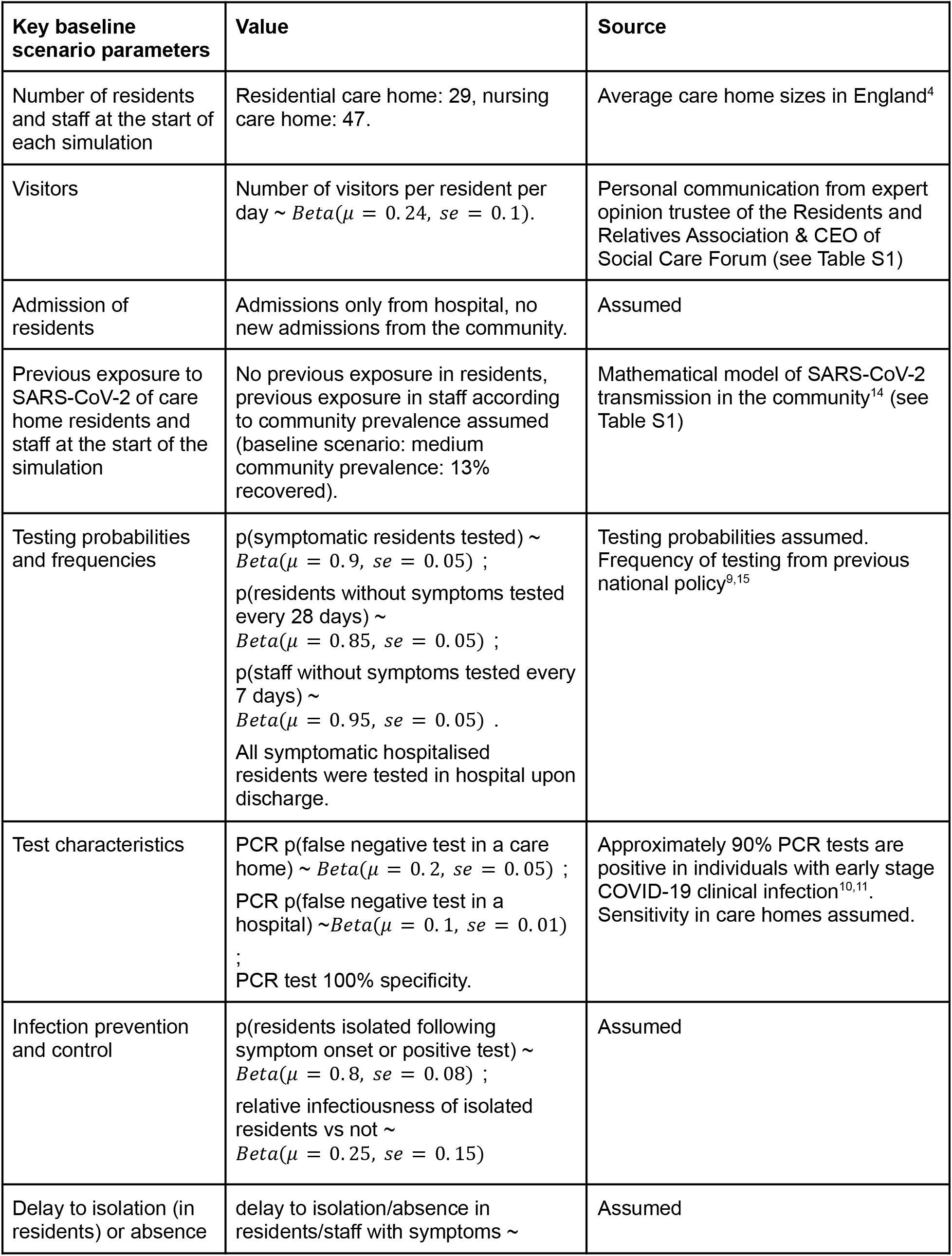
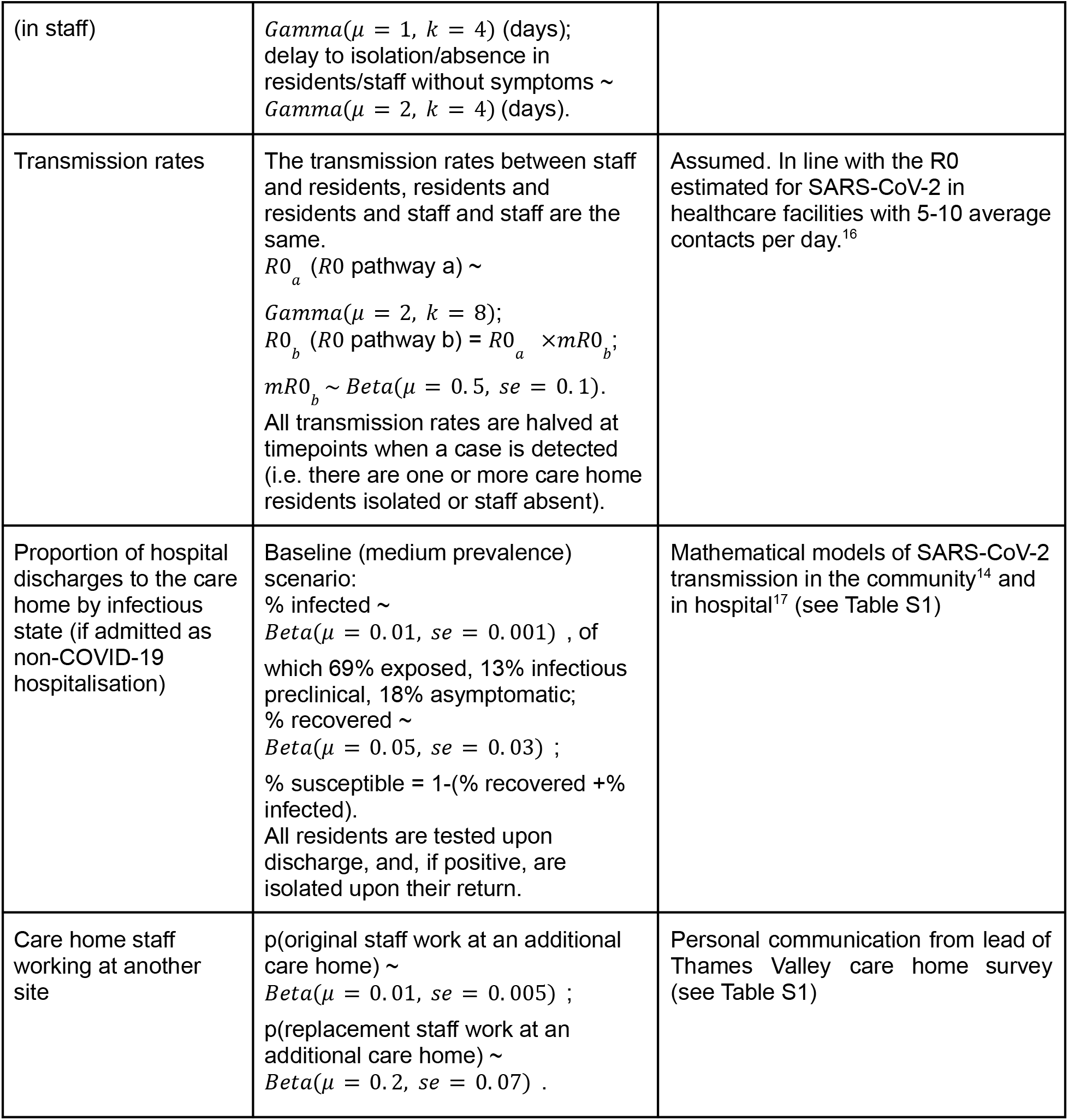
Key parameter values in the baseline scenario.

To assess the relative importance of the routes of entry into the care home, we evaluated four scenarios: eliminating the importation from hospital into the care home, stopping visitors, restricting all original staff to work in one care home, and stopping the importation from staff. To assess the impact of different testing strategies in care homes we varied the frequency of testing, compared PCR and LFD tests, and compared the testing of residents, staff, and both. We also simulated reactive improvements in IPC (modelled through a further reduction in the mean transmissibility in the care home once an outbreak is detected from 50% (95% 31-73%) to 75% (95% 63-85%)), decreasing the delay to isolation/absence to a mean of 0.25 days (95% 0.08-0.48) for residents/staff testing positive or being symptomatic, increasing the mean proportion of residents symptomatic and testing positive being isolated to 95% (95% 76-100%) and the mean effectiveness of isolation to 95% (65-100%), stopping all visiting, and restricting all original staff to work in one care home. The effectiveness of each intervention was calculated as described in Supplementary material A4.

Univariate sensitivity analyses were conducted for all parameters (see Supplementary material A1). We also estimated the outcomes for *R*0_*a*_ (*R*0 for the symptomatic pathway (a)) of 1 and 3.

These are in line with the R0 estimated for SARS-CoV-2 in healthcare facilities with 5-10 average contacts per day.^16^

### Shiny application COS-LTCF

The transmission model has been developed into a freely-available Shiny application found here: https://cmmid-lshtm.shinyapps.io/cos-ltcf/. COS-LTCF enables the user to explore alternative care home characteristics, outbreak characteristics and interventions to those considered here.

## Results

### Outbreak risk and role of community prevalence in driving outbreaks

Under baseline assumptions (Table 1), COVID-19 outbreaks were probable in both residential and nursing care homes despite the testing strategies and non-pharmaceutical interventions already in place (see Figures S6-S13 for outbreak dynamics in patients and staff). The median behaviour of the model predicts an outbreak in which approximately 40% of residential care home residents become infected and recover by day 90 (35% in nursing care homes), with a cumulative median of two deaths due to COVID-19 (none in residential care homes).

The probability of a care home experiencing an outbreak varied greatly depending on the community prevalence assumed (Figure 2). By day 30, a cumulative 10% (95% 4-27%) of nursing care homes had experienced outbreaks under low community prevalence, 42% (95% 16-83%) under medium prevalence, and 90% (95% 48-100%) under high prevalence. Outbreaks in residential care homes were somewhat less likely to occur by day 30 (respectively, 5%, 95% 2-14%, 23%, 95% 9-54%, 72%, 95% 37-98% by day 30). By day 90, a cumulative 24% (95% 1-65%) of nursing care homes had experienced large outbreaks under low community prevalence, 65% (95% 2-99%) under medium prevalence, and 89% (95% 8-100%) under high prevalence (in residential care homes, respectively, 6%, 95% 0-27%, 19%, 95% 1-79%, 42%, 95% 2-93%). In univariate sensitivity analyses, these outcomes were generally most sensitive to changes to assumed transmission rates and durations of infectiousness (Figures S14, S15). Overall, simulations are consistent with the dynamics observed in England, in which care homes are experiencing outbreaks despite the interventions in place.^3^

**Figure 2.**
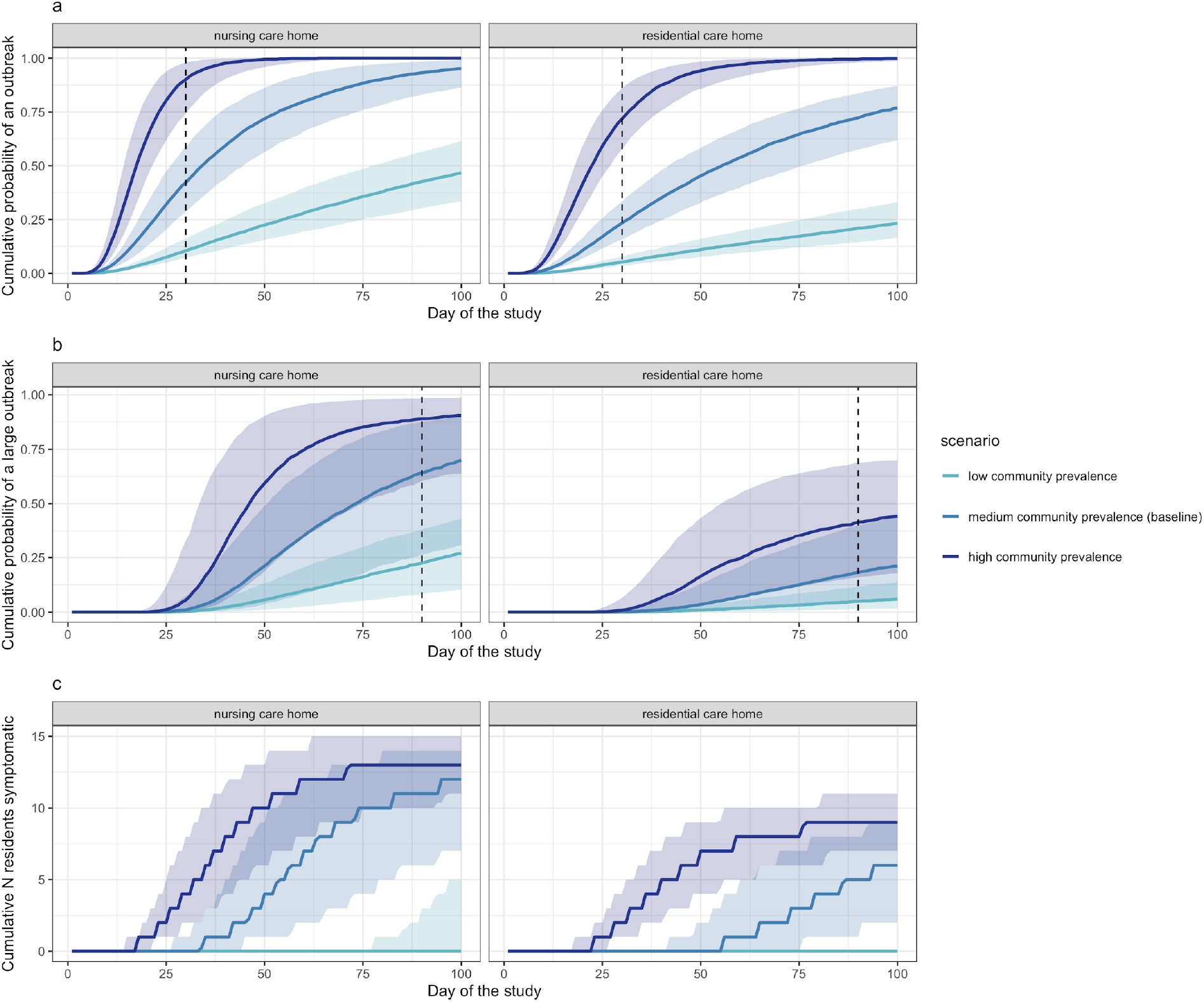
The cumulative probability of an outbreak (a), large outbreak (b) and number of residents symptomatic (c) over time is dependent on community prevalence (high = dark blue, medium (baseline) = light blue, low = turquoise) in both nursing care homes (left panels) and residential care homes (right panels). The coloured line represents the median and the shaded area represents the 25-75%. The vertical dashed lines show the thresholds (30 and 90 days) at which the cumulative probability of an outbreak and a large outbreak (respectively) were assessed in subsequent analysis.

### Routes of importation to the care home

We considered three routes of importation to the care home: from the community through staff, from the community through visitors, and from hospital through residents. We assumed staff had the same risk of acquiring SARS-CoV-2 outside of the care home as any other individual in the community, (except those working at more than one care home who had an additional risk). In the baseline scenario, each resident visited hospital on average 0.5 times per year (details in Supplementary material A2) and received 88 visitors per year (see Table S1). Compared to the baseline scenario, where all of these routes of importation are included, eliminating importation from hospital into the care home and stopping visitors had a small impact on the probability of an outbreak at 30 days (Figure 3). The most influential route of importation was through staff from the community. This ranking is robust to different community prevalence scenarios and across both types of care homes considered.

**Figure 3.**
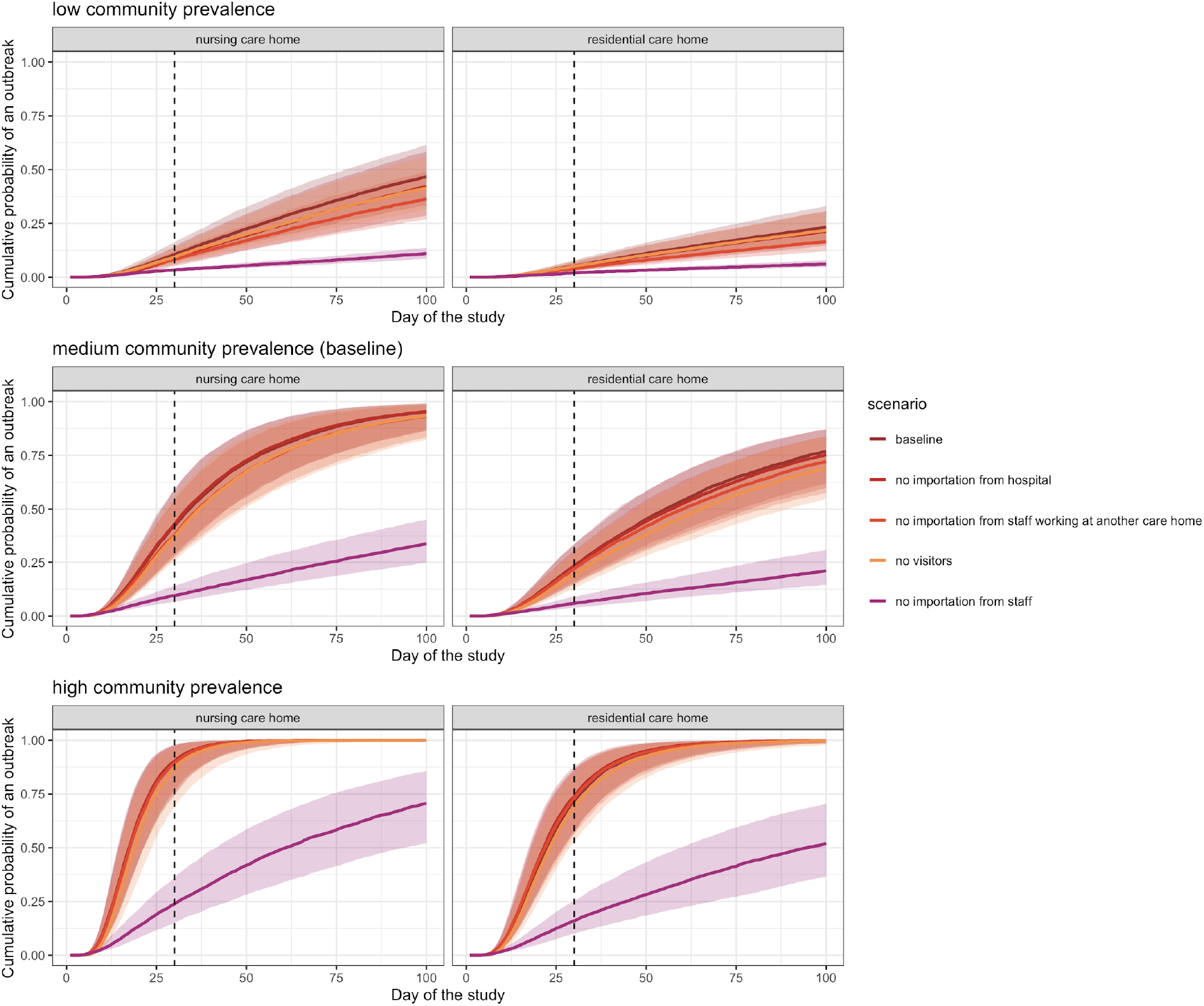
The cumulative probability of an outbreak at 30 days under low community prevalence (top panels), medium community prevalence (middle panels) and high community prevalence (bottom panels) over time for different importation scenarios (dark brown=baseline, dark red=no importation from hospital, light red=no importation from staff working at another care home, orange=no visitors, purple=no importation from staff), in both nursing care homes (left panels) and residential care homes (right panels).

### Testing strategy

Under baseline assumptions, in nursing care homes, the most effective testing strategy of those explored in reducing the cumulative probability of an outbreak at 30 days was daily LFD testing in staff and in residents (Figure 4). This strategy was 42% (95% 21-58%) effective in preventing outbreaks at 30 days compared to no testing under otherwise baseline scenario assumptions (i.e. it reduced the relative probability of an outbreak at 30 days by 42% compared to no testing). Eliminating testing in residents altogether when staff were tested daily by LFD yielded similar results (39% effective, 95% 18-55%, respectively). The effectiveness of PCR testing strategies was lower than for equivalent frequency LFD strategies.

**Figure 4.**
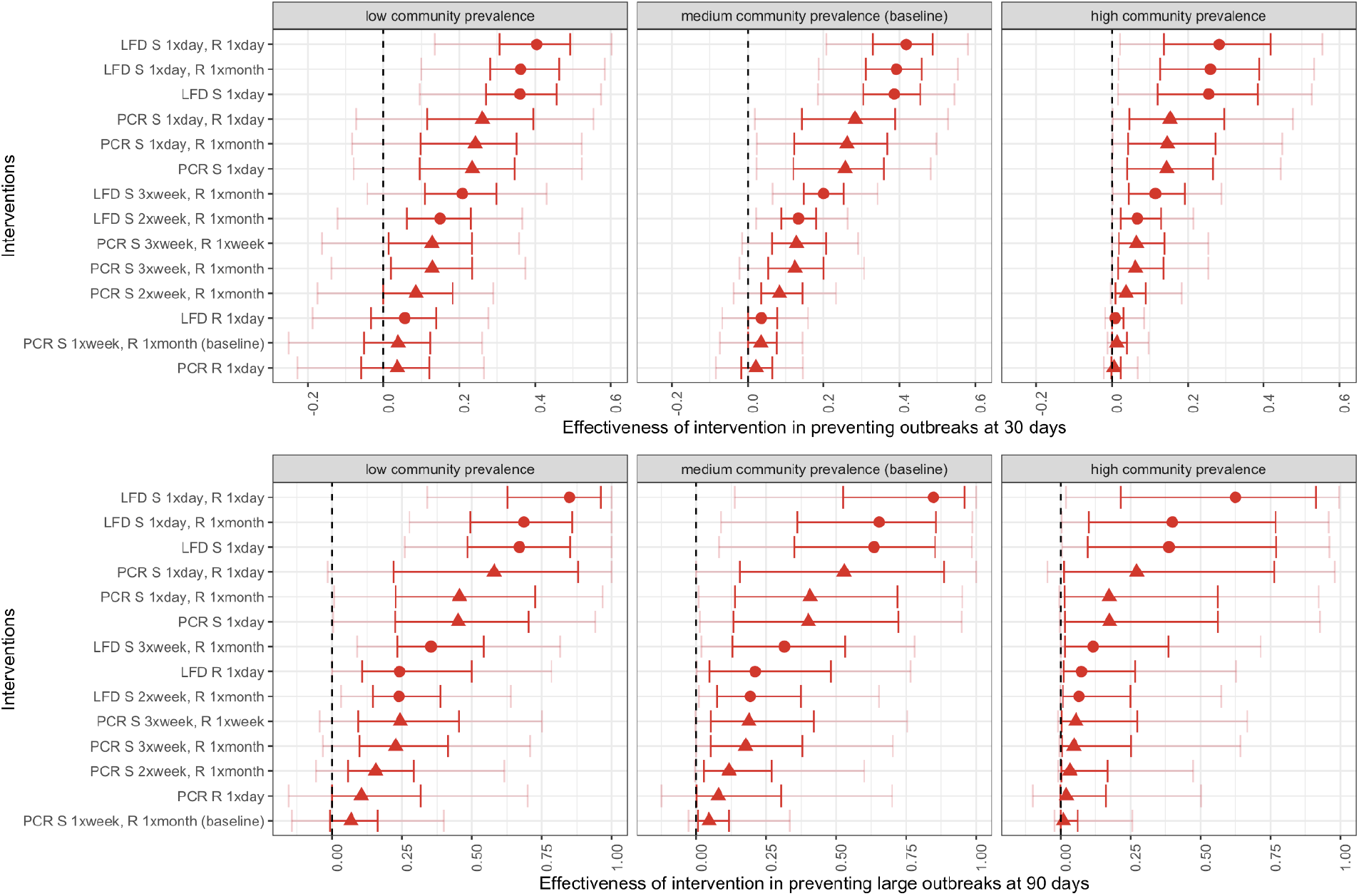
Effectiveness of testing strategies in preventing outbreaks in nursing care homes at 30 days (top panels) and large outbreaks at 90 days (bottom panels) by testing intervention and and under low (left panels), medium (baseline, middle panels) and high (right panels) community prevalence. In red, the 25-75%, in pink, the 5-95%. Testing interventions include PCR testing (triangles) and LFD testing (dots). R stands for resident and S for staff.

**Figure 5.**
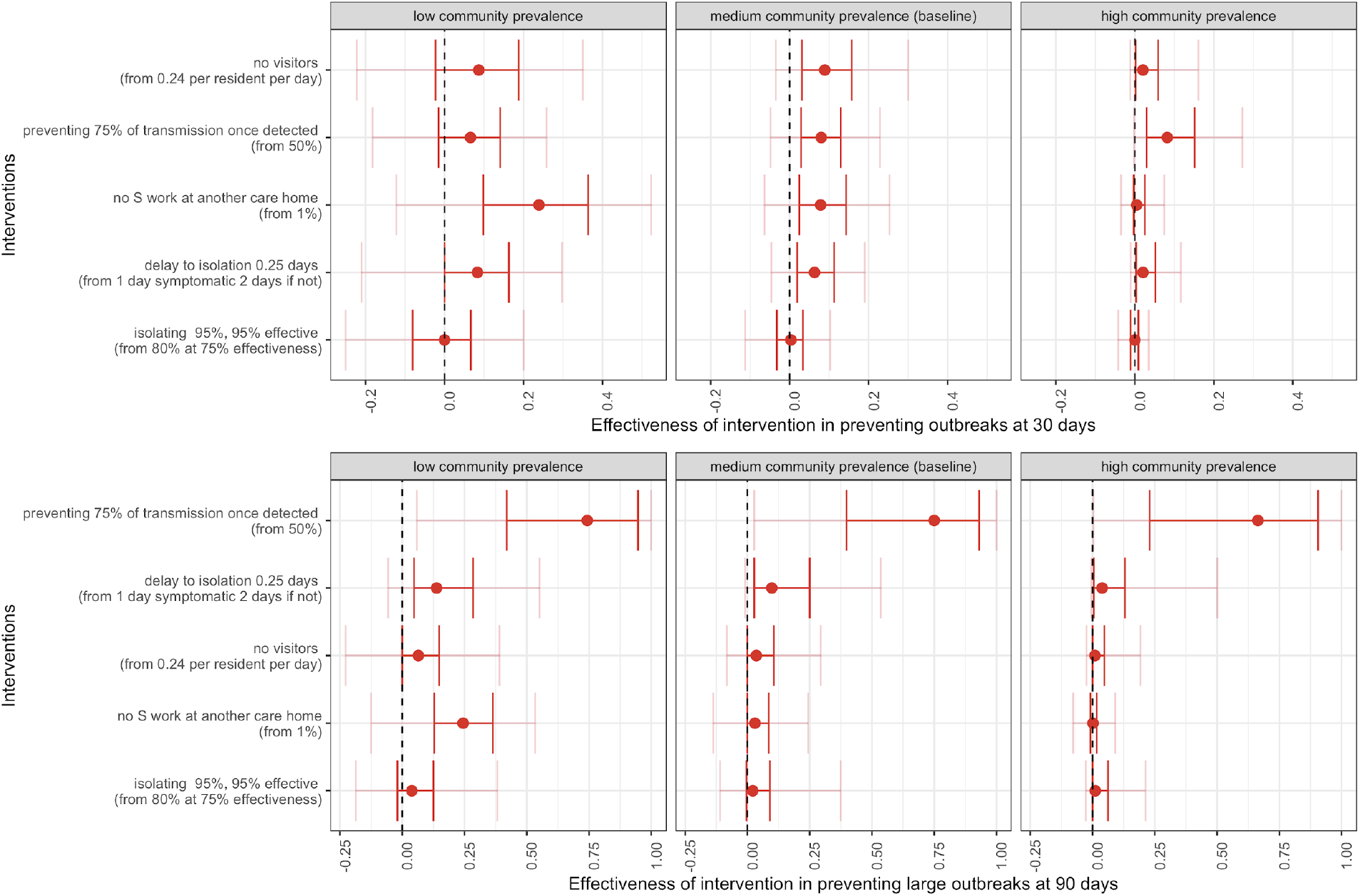
Effectiveness of interventions in preventing outbreaks in nursing care homes at 30 days (top panels) and large outbreaks at 90 days (bottom panels) by low (left panels), medium (baseline, middle panels) and high (right panels) community prevalence. In red, the 25-75%, in pink, the 5-95%. R stands for resident and S for staff.

Testing only residents daily showed no significant efficacy (2%, 95% −8-14%, by PCR and 4%, 95% −7-16% by LFD). The impact of the baseline testing strategy (PCR once a week in staff, and once a month in residents) compared to no testing was also negligible (3%, 95% −7-14%). The effect of increasing the frequency of testing in staff to twice or three times a week was small (8% effective, 95% −4-23%, and 12% effective, 95% −2-31%, respectively by PCR and 13% effective, 95% 2-26%, and 20% effective, 95% 7-34%, respectively by LFD).

Similar patterns of testing effectiveness were observed in residential care homes (see Figure S16). Assuming a higher mean R0 for pathway (a) reduced the effectiveness of all testing strategies considered (see Figure S17).

### Other IPC strategies

Under medium community prevalence, reactive improvements in IPC in nursing care homes (decreasing the transmission rate by 75% vs. 50% once an outbreak was detected) were 75% effective (95% 3-100%) in averting large outbreaks at 90 days compared to baseline measures (See Figure S17 for residential care homes, and S18 for different *R*0_*a*_ assumptions). Proactive improvements in IPC are also important, as shown by the differences in outcome estimated for different assumptions of R0. When the mean R0 in pathway (a) was decreased from 2 to 1, the probability of an outbreak at 30 days decreased from 42% (95% 16-83%) to 22% (95% 8-58%) and the probability of a large outbreak at 90 days from 65% (95% 2-99%) to 8% (95%0-72%).

Decreasing the delay to isolation or absence to a mean of 0.25 days for residents or staff testing positive or symptomatic (from a mean of 1 and 2 days, respectively, in the baseline scenario) was 10% effective (95% −1-54%) in averting large outbreaks at 90 days. Increasing the mean proportion of symptomatic and test positive residents being isolated to 95% (from a mean of 80%) and the mean effectiveness of isolation to 95% (from a mean of 75%), restricting all visitors to the care home (from a mean frequency of 0.24 per resident per day), and limiting all original staff to working in one care home (from a mean of 1% working at more than one site) had a small effectiveness in averting large outbreaks at 90 days.

## Discussion

Our study shows that COVID-19 outbreaks are probable in both residential and nursing care homes in England despite the non-pharmaceutical interventions in place, suggesting that additional measures are necessary to prevent and contain outbreaks in this setting. This is consistent with COVID-19 deaths in care homes rising during the third wave of the pandemic.^3^ We show that community prevalence, through staff importation, determines to a large extent the probability of outbreaks at 30 days. Importation through visitors at pre-pandemic levels and through the infection of residents during a hospital admission (after which all residents are now tested), are less likely to cause outbreaks. This is in agreement with a recent study in Wales finding that the risk of care home outbreaks was not significantly increased in the period following a hospital discharge to the care home.^18^ These findings suggest that reducing importations via care home staff should be the main focus of interventions aiming to prevent the importation of SARS-CoV-2 to care homes.

The most effective testing strategies involve daily testing of staff. Whilst some of the testing capability limitations in England could be addressed simply, others, such as the time pressure on staff (who are already often overstretched) to carry out additional tests remain problematic. We find that testing strategies involving only residents are ineffective in preventing outbreaks. LFD testing had a marginal benefit over PCR due to the lower delay of turnaround of the test, despite poorer sensitivity. However, we did not account for false negative tests contributing to a false sense of security that could lead to increased transmission. Our qualitative findings on the frequency, type of test and the best population to test are in line with those from recent mathematical models describing SARS-CoV-2 transmission in care homes examining testing strategies in other countries.^19–23^ However, the particular testing frequency needed to substantially reduce the probability of an outbreak is context-specific, and heavily dependent on the modelling assumptions made (e.g. baseline considered, contact rates assumed, infectious period, proportion of staff and residents asymptomatic, delay to isolation).

We assessed the effectiveness of other IPC interventions when added to the current strategies in place (baseline). We show that decreasing transmission rates within the care home, whether proactively or reactively, was very effective in averting outbreaks at 30 days and large outbreaks at 90 days. Further research is needed to quantify the single and combined effect of measures such as deep cleaning, PPE, enhanced hand hygiene, ventilation and cohorting of residents and staff on decreasing transmission rates in this setting.

Our findings suggest that when 1% of staff work at more than one care home, limiting all original staff to working at one site is ineffective; however, this may be useful for care homes with a higher proportion of staff working across various sites, as SARS-CoV-2 has been shown to spread between care homes. Although the proportion of care homes with staff working at multiple care homes has been described^5^, only one survey of Thames Valley care home managers has estimated the proportion of staff working across multiple care homes (1%, pers. comm. Dr Watson June 2020). Staff constitute an important route of importation into care homes, therefore, it is necessary to better understand their working patterns and behaviours.

We found restricting visitors in English care homes was also ineffective. A recent rapid review of the literature found no evidence of an impact of visitors on COVID-19 infections in care homes, but an increase in depression, loneliness and a potential impact on the quality of care of residents due to the absence of informal care.^24^ A mathematical modelling study of Scotish care homes also found the impact of visiting on COVID-19 outbreaks to be negligible.^19^ Together, these findings suggest visiting restrictions may provide more harm than benefit to residents, provided baseline IPC measures are in place. However; to our knowledge, no studies to date have published visiting patterns for care homes in England. In addition to relatives, professionals also visit care home residents (e.g. GPs, physiotherapists). We did not account for these visits in our model and may therefore underestimate the overall frequency and impact of visits. We also did not account for the uniqueness and correlation between visitors.

Another key data limitation is the need to better understand the contact patterns within care homes. We assumed that transmission rates were the same between residents, between residents and staff, and between staff. Contrasting our findings, a recent study set in a rehabilitation centre where the contacts between residents were common and prolonged found that testing residents was more effective than testing staff.^25^ This shows the importance that contact matrices may have on determining appropriate interventions in care homes. We also assumed transmission rates were the same in nursing and residential care homes; however, nursing care homes may have more staff-resident contact due to the higher care needs of this population, whilst resident-resident contacts may be lower. Another limitation of this work is that we do not consider the effect of staff absence on the rates of transmission within the care home, which are likely to increase due to remaining staff being overstretched and therefore more likely to carry out sub-standard IPC.

To the best of our knowledge, this is the first model to explicitly evaluate the relative importance of all SARS-CoV-2 importation routes to care homes, including the hospitalisation of residents, and the first study to assess the impact of a range of non-pharmaceutical interventions against SARS-CoV-2 in English care homes. We also developed the COS-LTCF app to enable decision makers to explicitly tailor the care home, outbreak and intervention characteristics to their particular setting of interest. Our study highlights the high risk of a COVID-19 outbreak occurring English care homes under baseline IPC interventions. Our model indicates that community prevalence, through staff importation, is key in determining the probability of care home outbreaks at 30 days, and that more frequent testing is needed. Our preliminary analysis shows that when 50% of residents and staff were immune at the start of the simulations, 57% (95% 42-62%) of outbreaks and 99% (95% 69-100%) of large outbreaks were averted in nursing care homes, compared to baseline, which is the most effective intervention considered. This suggests that vaccination, even if only partially effective, will provide a substantial effect in reducing the burden of disease in care homes. Future work will explore the dynamics of vaccination in this setting.

## Data Availability

Available upon request.

## Contributors

WJE and AR designed the study and the model. AR led the development and analysis of the transmission model, and writing of the manuscript. WJE, RCB, DRMS, GMK, and The Centre for the Mathematical Modelling of Infectious Diseases COVID-19 working group contributed to shaping the analysis and writing the manuscript. NGD and SE contributed outputs from the community and hospital transmission models they respectively developed, FG and SRD led the analysis of SUS data and contributed to writing the manuscript. AR and RCB developed the Shiny application. All authors read and approved the final report.

## Acknowledgements

AR acknowledges funding from the National Institute for Health Research (grant: PR-OD-1017-20002). RCB acknowledges funding from the European Union Horizon 2020 Research and Innovation programme, project EpiPose (101003688), throughout the duration of the study. DS is supported by a Canadian Institutes of Health Research Doctoral Foreign Study Award (Funding Reference Number 164263) and the French National Research Agency project SPHINX-17-CE36-0008-01. GMK is supported by the UK Medical Research Council (grant: MR/P014658/1). We would like to thank Ian Hall, as well as the rest of the Care Home Working Group for helpful conversations as well as William Laing, Claire Goodman, Des Kelly and Julie Robotham for their insight; and Carl Pearson and Joel Hellewell for their helpful inputs. This work uses data provided by patients and collected by the NHS as part of their care and support. The development of the Shiny application COS-LTCF was funded by the World Health Organization.

