## Supplementary material for "Impact of non-pharmaceutical interventions on SARS-CoV-2 outbreaks in English care homes: a modelling study"

### Appendix

**Supplementary tables -- p 1**
**Supplementary figures -- p 10**
**Supplementary material A0 -- p 28**
**Supplementary material A1 -- p 29**
**Supplementary material A2 -- p 35**
**Supplementary material A3 -- p 39**
**Supplementary material A3 -- p 41**
**References appendix -- p 42**

**SUPPLEMENTARY TABLES**

*Table S1. Parameter values and sources*

| Parameter | Description | Value / distribution | Source |
|---|---|---|---|
| Parameters relating to the SARS-CoV-2 infection process | | | |
| $1/\boldsymbol{\varepsilon}$ | Duration of latent period (days) | ~ $Gamma(\mu = 2.5,\ k = 4)$ | Incubation period (latent period plus duration of preclinical infection) of mean 5 days[1] subdivided into latent period (mean 2.5) and preclinical infection (mean 2.5 days). |
| $1/\tau$ or *d_preclinical_infection* | Duration of preclinical infection (days) | ~ $Gamma(\mu = 2.5,\ k = 4)$ | Incubation period (latent period plus duration of preclinical infection) of mean 5 days[1] subdivided into latent period (mean 2.5) and preclinical infection (mean 2.5 days). |
| *d_clinical_infection* ($1/\omega$+$1/\varphi$) | Duration of clinical infection (symptomatic disease) (days) | ~ $Gamma(\mu = 5,\ k = 4)$ | Assumed duration of clinical infection of mean 5 days (approximated from[2–4]), subdivided into highly infectious clinical infection and less infectious clinical infection, corresponding to a linear decrease in viral load.[2] |
| $1/\omega$or *d_clinical_infection_h* | Duration of early highly infectious stage of clinical infection (days) | *d_clinical_infection/2* | |
| $1/\varphi$ or *d_clinical_infection_l* | Duration of late low infectious stage of clinical infection (days) | *d_clinical_infection/2* | |
| *d_total_infectiousness* or *d_asymptomatic_infection* or $1/\varphi_a$ | Total duration of infectiousness or duration of asymptomatic infection (days) | $1/\tau\ +\ 1/\omega\ +\ 1/\varphi$ | |

| | | | |
|---|---|---|---|
| $R0_a$ | R0 in the care home for pathway (a), ie. the transmission pathway in which individuals (eventually) present with symptoms | Baseline: ~ $Gamma(\mu = 2, k = 8)$ | Assumed (baseline). |
| $p_{transmission\_pc\_vs\_c}$ | Probability that transmission in pathway (a) occurs during the preclinical stage compared to the clinical stages | ~ $Beta(\mu = 0.4, se = 0.05$ | Pre-clinical stage on average 40% of transmission compared to clinical stage.[2,3] |
| $p_{transmission\_cl\_vs\_ch}$ | Probability that transmission during the clinical stages occurs during the late low infectiousness stage (a) compared to the early high infectiousness stage | ~ $Beta(\mu = 0.3, se = 0.05$ | We assumed a linear decrease in viral load over the symptomatic period.[2] |
| $m_{R0_b}$ | Relative R0 pathway (b) vs pathway (a) | ~ $Beta(\mu = 0.5, se = 0.1)$ | We assumed the relative infectiousness of pathway (b) was on average half of that of (a).[5] |
| $m\beta_d$ | Relative transmission rates in care home when an infected resident/staff member is detected (ie. ≥1 resident isolated or staff member absent) vs not | Baseline: ~ $Gamma(\mu = 0.5, k = 15$ | Assumed (baseline). |
| $m_i$ | Relative infectiousness of residents being isolated vs not | ~ $Beta(\mu = 0.25, se = 0.1$ | Assumed (baseline). |
| $n\_V$ | Number of visitors per resident in the care home on any one day | Baseline: ~ $Beta(\mu = 0.24, se = 0.1$ | Baseline: expert opinion trustee of the Residents and Relatives Association & CEO of Social Care |

| | | | |
|---|---|---|---|
| | | | Forum personal communication ("around 10% don't have any and 10% have visitors every day. Of the remaining 80% I reckon they could be divided between half having a couple of visitors a week and half 1 visitor every 2 weeks"). (0.1*1)+(0.4*(2/7))+(0.4*(1/14))= 0.24 |
| $p_{V_{infectious}}$ | Probability of a visitor being infectious with SARS-CoV-2 | Baseline (medium community prevalence scenario): $\sim Gamma(\mu = 0.002, k = 20)$; low community prevalence scenario: $\sim Gamma(\mu = 0.0004, k = 20)$ high community prevalence scenario: $\sim Gamma(\mu = 0.01, k = 20)$ | Median of 500 runs p(infected with preclinical infection)+p(infected asymptomatic) in England from Davies et al. 2020 model[5] on 2020-09-30 (baseline, medium community prevalence scenario), 2020-07-15 (low community prevalence scenario) or 2020-04-01 (high community prevalence scenario). |
| $p_a$ | Probability an exposed resident becomes asymptomatic | $\sim Beta(\mu = 0.4, se = 0.1)$ | The proportion of infected residents with asymptomatic infection ranged from 6-45% in the literature .[6–11] |
| $p_{as}$ | Probability an exposed staff member becomes asymptomatic | $\sim Beta(\mu = 0.5, se = 0.05$ [illegible] | Assumed.[5] |
| $R_C$ | Reproduction number in the community | Baseline (medium community prevalence scenario): $\sim Gamma(\mu = 1, k = 16)$; low community prevalence scenario: $\sim Gamma(\mu = 0.8, k = 16)$; | Assumed. |

| | | | |
|---|---|---|---|
| | | high community prevalence scenario: ~ $Gamma(\mu = 1.5, k = 16)$ | |
| $d\_total\_infectiousness\_C$ | Duration of infectiousness in the community (days) | ~ $Gamma(\mu = 5, k = 4)$ | Assumed 5 days.[5] |
| $p_{C_{infectious}}$ | Probability of a community member being infectious with SARS-CoV-2 | Baseline(medium community prevalence scenario): ~ $Gamma(\mu = 0.003, k = 20)$; low community prevalence scenario: ~ $Gamma(\mu = 0.0005, k = 20)$; high community prevalence scenario: ~ $Gamma(\mu = 0.01, k = 20)$; | Median of 500 runs p(infected with preclinical infection)+p(infected asymptomatic)+p(infected clinical infection) in England from Davies et al. 2020 model[5] on the 2020-09-30 (baseline, medium community prevalence scenario), 2020-07-15 (low community prevalence scenario) or 2020-04-01 (high community prevalence scenario). |
| $p_{original\ staff\ \ work\ at\ care\ home\ 2}$ | Probability of original staff working at another care home | ~$Beta(\mu = 0.01, se = 0.005)$ | Pers. comm. lead of care home survey Thames Valley, Dr Conall Watson June 2020. |
| $p_{replacement\ staff\ \ work\ at\ care\ hom}$ [illegible] | Probability of replacement staff working at another care home | ~ $Beta(\mu = 0.2, se = 0.0$ [illegible] | Assumed. |
| $p_{care\ home\ outbreak}$ | Probability that another care home is experiencing an outbreak | Baseline (medium community prevalence scenario): ~ $Beta(\mu = 0.03, se = 0.$ [illegible] ; low community prevalence scenario :~ $Beta(\mu = 0.01, se = 0.$ [illegible] ; high community prevalence scenario :~ $Beta(\mu = 0.1, se = 0.0$ [illegible] | Assuming 25% of care homes have an ongoing outbreak, i.e. ~4,000 of 15,000 care homes in baseline scenario (unpublished data, Public Health England). In the high community prevalence scenario we assumed 40% of care homes could have an outbreak and in the low community |

| | | | |
|---|---|---|---|
| | | | prevalence scenario, 10%. |
| $p_{res\ inf}$or $p_{staff\ inf}$ | Probability that a resident in a care home with an outbreak is infectious or probability that a staff member in a care home with an outbreak is infectious | ~ $Beta(\mu = 0.1,\ se = 0.0$[illegible] | Corresponds to approximately 3 residents or staff in an average residential care home being infectious. |
| $1/\nu$ | Duration of staff absence (days) | ~ $Gamma(\mu = 14,$ $k = 40)$ | Assuming staff absent for 14 days. |
| Parameters relating to flows in and out of the care home | | | |
| $\delta$ | Death rate pppd in the care home for residents without COVID-19 clinical infection | ~ $Gamma(\mu = 0.0005,$ $k = 12)$for residential care homes and ~ $Gamma(\mu = 0.001,$ $k = 12)$for nursing care homes | See Supplementary material A1. |
| *CFR* | Case fatality ratio in residents | ~ $Beta(\mu = 0.25,\ se = 0.$[illegible] | Literature estimates range from 17-36%[6–9,12,13] |
| $p_{death\ res\ within\ care\ home}$ | Probability resident with COVID-19 dies within the care home (vs. hospital) | ~ $Beta(\mu = 0.8,\ se = 0.0$[illegible] | 80% of these deaths assumed to take place in the care home (the remaining 20% in hospital).[14] |
| $m_{\delta rvn}$ | Relative death rate in residential vs nursing care homes | ~ $Beta(\mu = 0.47,\ se = 0.$[illegible] | See Supplementary material A3. |
| $\kappa$ | Hospitalisation rate pppd for residents without COVID-19 clinical infection | Baseline (medium community prevalence scenario): ~ $Gamma(\mu = 0.00132,$ $k = 10)$; low community prevalence scenario: ~ $Gamma(\mu = 0.00130,$ $k = 10)$; high community prevalence scenario: ~ $Gamma(\mu = 0.00104,$ $k = 10)$ | See Supplementary material A1. |
| $p_{survival}$ | Probability of residents with | ~ $Beta(\mu = 0.7,\ se = 0.1$[illegible] | SUS data showed a mean of 30% of |

| | | | |
|---|---|---|---|
| | COVID-19 clinical infection returning to the care home from hospital having survived | | patients admitted to hospital for COVID died in hospital (see Supplementary material A4 for details). |
| $p_{EI\ non-COVID-19\ H}$ | Proportion of hospital discharges that enter the care home infected states (E, Ipc, Ia) (residents that didn't go to hospital symptomatic for COVID-19) | Baseline (medium community prevalence scenario): ~ $Beta(\mu = 0.01, se = 0.001)$ ; low community prevalence scenario: ~ $Beta(\mu = 0.007, se = 0.001)$ ; high community prevalence scenario: ~ $Beta(\mu = 0.004, se = 0.001)$ | Mean prevalence in each of the model compartments exiting hospital from the Evans et al. 2020 mathematical model on the 2020-05-01 (baseline, medium community prevalence scenario), 2020-07-15 (low community prevalence scenario) or 2020-04-01 (high community prevalence scenario). [15] |
| $p_{E,EI\ non-COVID-19\ H}$, $p_{Ipc,EI\ non-COVID-19\ H}$, $p_{Ia,EI\ non-COVID-19\ H}$ | Proportion of infected hospital discharges that enter the care home exposed (E), infectious preclinical (Ipc), and asymptomatic (Ia) states (residents that didn't go to hospital symptomatic for COVID-19) | Baseline (medium community prevalence scenario): 69% exposed, 13% infectious pre-clinical, 18% asymptomatic; low community prevalence scenario: 59% exposed, 14% infectious pre-clinical, 27% asymptomatic; high community prevalence scenario: 74% exposed, 11% infectious pre-clinical, 15% asymptomatic. | Mean prevalence in each of the model compartments exiting hospital from the Evans et al. 2020 mathematical model on the 2020-05-01 (baseline, medium community prevalence scenario), 2020-07-15 (low community prevalence scenario) or 2020-04-01 (high community prevalence scenario). [15] |
| $p_{R\ non-COVID-19\ H}$ | Proportion of hospital discharges that enter the care home recovered (R) state (residents that didn't go to hospital symptomatic for COVID-19) | Baseline (medium community prevalence scenario): ~ $Beta(\mu = 0.05, se = 0.$[illegible] ; low community prevalence scenario: ~ $Beta(\mu = 0.06, se = 0.$[illegible] ; high community prevalence scenario: ~ $Beta(\mu = 0.02, se = 0.$[illegible] | Assuming the mean ratio of susceptible to recovered was the same as in the community (informed by median of 500 runs in England from Davies et al. 2020 model fit to data from the community[5]). This ratio was applied to $(1 - p_{I\ non-COVID-19\ H})$, estimated as above. |

| | | | |
|---|---|---|---|
| $p_{S\ non-COVID-19\ H}$ | Proportion of hospital discharges that enter the care home susceptible (S) state (residents that didn't go to hospital symptomatic for COVID-19) | $1 - (p_{R\ non-COVID-19\ H} + p_{EI\ non-COVID-19\ H})$ | |
| $p_{Icl\ COVID-19\ H}$ | Proportion of hospital discharged residents that enter the care home still residually shedding ie. in state of clinical infection with low infectiousness (Icl) vs recovered (R) state (residents that went to hospital symptomatic for COVID-19) | ~ $Beta(\mu = 0.06, se = 0.0$ | Assuming on average 6% were still shedding at 14 days[4], when they were assumed to (on average) be discharged. In the baseline scenario, those testing positive upon hospital discharge test were immediately isolated upon their return to the care home (Icli compartment). |
| $p_{EI\ s2}$ | Proportion of replacement staff who enter the care home infected states (E, Ipc, Ia) | Baseline (medium community prevalence scenario): ~ $Beta(\mu = 0.005, se = 0.001)$ ; low community prevalence scenario: ~ $Beta(\mu = 0.0008, se = 0.001)$ ; high community prevalence scenario: ~ $Beta(\mu = 0.02, se = 0.001)$ | Assumed the same as in the community. Informed by median of 500 runs in England from Davies et al. 2020 model[5] on the 2020-09-30 (baseline, medium community prevalence scenario), 2020-07-15 (low community prevalence scenario) or 2020-04-01 (high community prevalence scenario). |
| $p_{E,\ EI\ s2}$, $p_{Ipc,\ EI\ s2}$, $p_{Ia,\ EI\ s2}$ | Proportion of infected replacement staff who enter the care home exposed (E), infectious preclinical (Ipc), and asymptomatic (Ia) states | Baseline (medium community prevalence scenario): 44% exposed, 12% infectious pre-clinical, 44% asymptomatic; low community prevalence scenario: 39% exposed, 15% infectious pre-clinical, 46% asymptomatic; high community | Assumed the same as in the community. Informed by median of 500 runs in England from Davies et al. 2020 model[5] on the 2020-09-30 (baseline, medium community prevalence scenario), 2020-07-15 (low community prevalence scenario) or |

| | | | |
|---|---|---|---|
| | | prevalence scenario: 38% exposed, 10% infectious pre-clinical, 52% asymptomatic. | 2020-04-01 (high community prevalence scenario). |
| $p_{R\,s2}$ | Proportion of replacement staff who enter the care home recovered (R) state | Baseline (medium community prevalence scenario): ~ $Beta(\mu = 0.13,\ se = 0.$[illegible] ; low community prevalence scenario: ~ $Beta(\mu = 0.11,\ se = 0.$[illegible] ; high community prevalence scenario: ~ $Beta(\mu = 0.04,\ se = 0.$[illegible] | Assumed the same as in the community. Informed by median of 500 runs in England from Davies et al. 2020 model[5] on the 2020-09-30 (baseline, medium community prevalence scenario), 2020-07-15 (low community prevalence scenario) or 2020-04-01 (high community prevalence scenario). |
| $p_{S\,s2}$ | Proportion of replacement staff who enter the care home susceptible (S) state | $1 - (p_{R\,s2} + p_{EI\,s2})$ | |
| Parameters relating to testing and isolation | | | |
| $1/\gamma_c$ or *delay_isolation_c* | Delay to isolation/absence in symptomatic residents/staff (days) | Baseline: $Gamma(\mu = 1, k = 4)$ | Assumed (baseline scenario). |
| $1/\gamma_{nc}$ or *delay_isolation_nc* | Delay to isolation/absence in residents/staff with preclinical infection or asymptomatic infection testing positive (days) | Baseline: $Gamma(\mu = 2, k = 4)$ | Assumed (baseline scenario). |
| $p_{tnc}$ | Probability of residents without COVID-19 clinical infection being tested | Baseline: ~ $Beta(\mu = 0.85,\ se = 0.$[illegible] | Assumed (baseline scenario). |
| $p_{tncs}$ | Probability of staff without COVID-19 clinical infection being tested | Baseline: ~ $Beta(\mu = 0.95,\ se = 0.$[illegible] | Assumed (baseline scenario). |
| $p_{tc}$ | Probability of residents with COVID-19 clinical | Baseline: ~ $Beta(\mu = 0.9,\ se = 0.05$[illegible] | Assumed (baseline scenario). |

| | | | |
|---|---|---|---|
| | infection being tested | | |
| $f_{tnc}$ | Frequency of testing residents without COVID-19 clinical infection (days) | Baseline: 28 | Assumed (baseline scenario). |
| $f_{tncs}$ | Frequency of testing staff without COVID-19 clinical infection (days) | Baseline: 7 | Assumed (baseline scenario). |
| $f_{tc}$ | Frequency of testing residents with COVID-19 clinical infection (days) | Baseline: 0 (testing on the day of symptom onset) | Assumed (baseline scenario). |
| $p_{fn}$ | Probability of false negative PCR result in care home | ~ $Beta(\mu = 0.2, se = 0.05$ | About 90% PCR tests are positive in individuals with early stage COVID-19 clinical infection.[16,17] However, assuming this is lower as staff are testing. |
| $p_{fnh}$ | Probability of false negative PCR result in hospital | ~ $Beta(\mu = 0.1, se = 0.01$ | About 90% PCR tests are positive in individuals with early stage COVID-19 clinical infection.[16,17] |
| $p_i$ | Probability of residents with COVID-19 clinical infection or a positive test being isolated | ~ $Beta(\mu = 0.8, se = 0.08$ | Assumed. |
| $p_{replaced}$ | Probability of absent staff being replaced | ~$Beta(\mu = 0.8, se = 0.05)$ | Assumed. |

Unless otherwise indicated, values assumed the same for nursing and residential care care homes.

**SUPPLEMENTARY FIGURES**

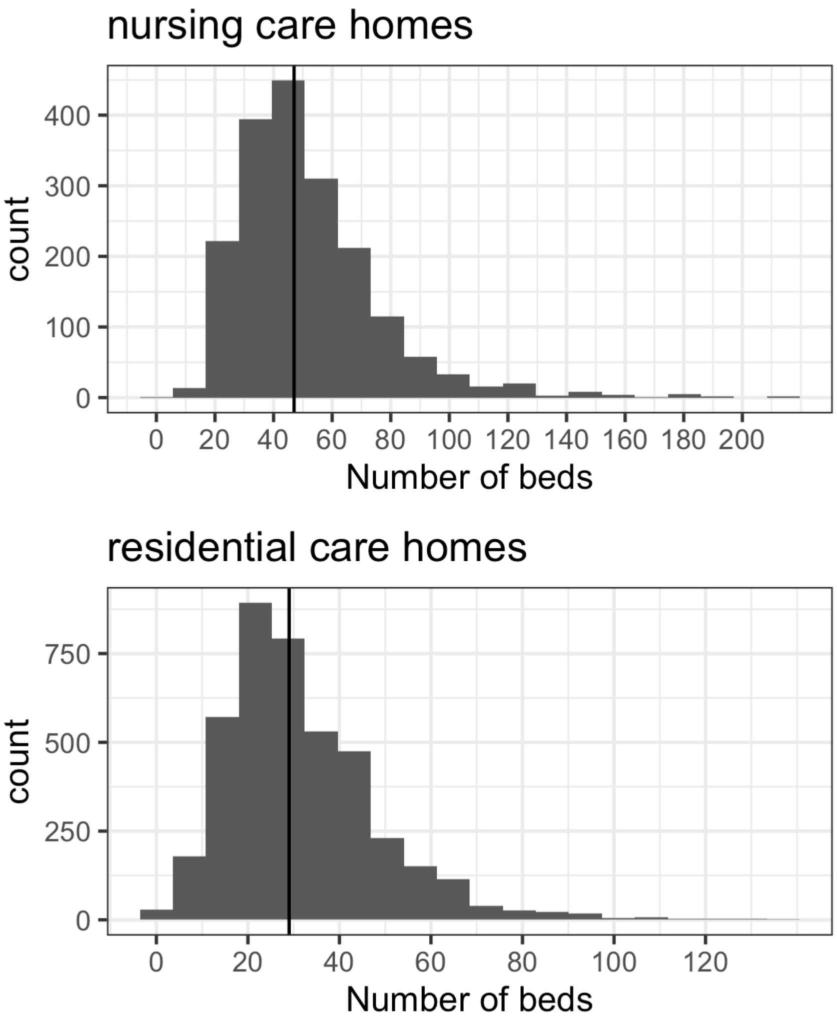


*Figure S1. Number of beds in nursing (top) and residential (bottom) care homes housing only older residents in England. Source: CQC database.*

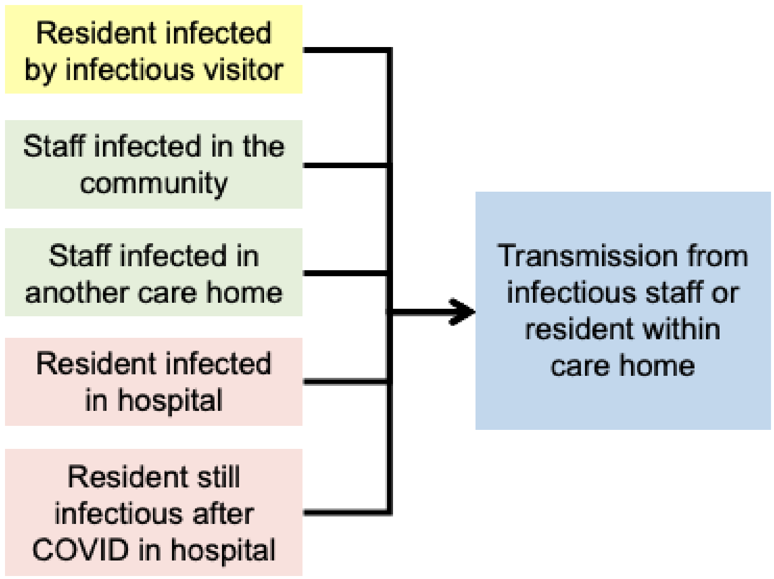


*Figure S2. Pathways by which care home residents and staff may become infected.*

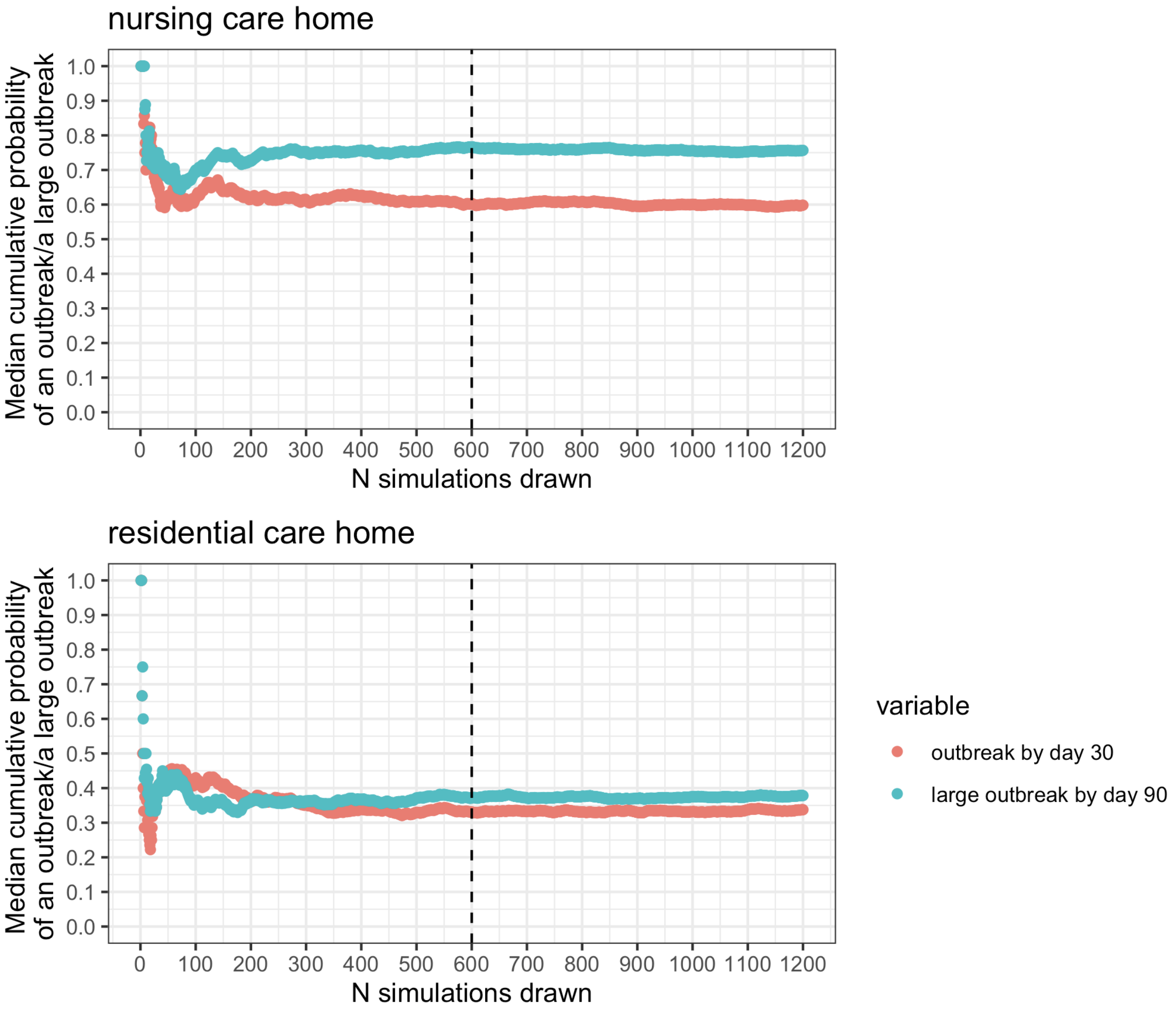


*Figure S3. Cumulative probability of an outbreak by day 30 (red) and a large outbreak by day 90 (blue) in a nursing care home (top plot) and a residential care home (bottom plot) by the number of simulations run for a single parameter set. The dashed line represents the number of simulations carried out for each parameter set.*

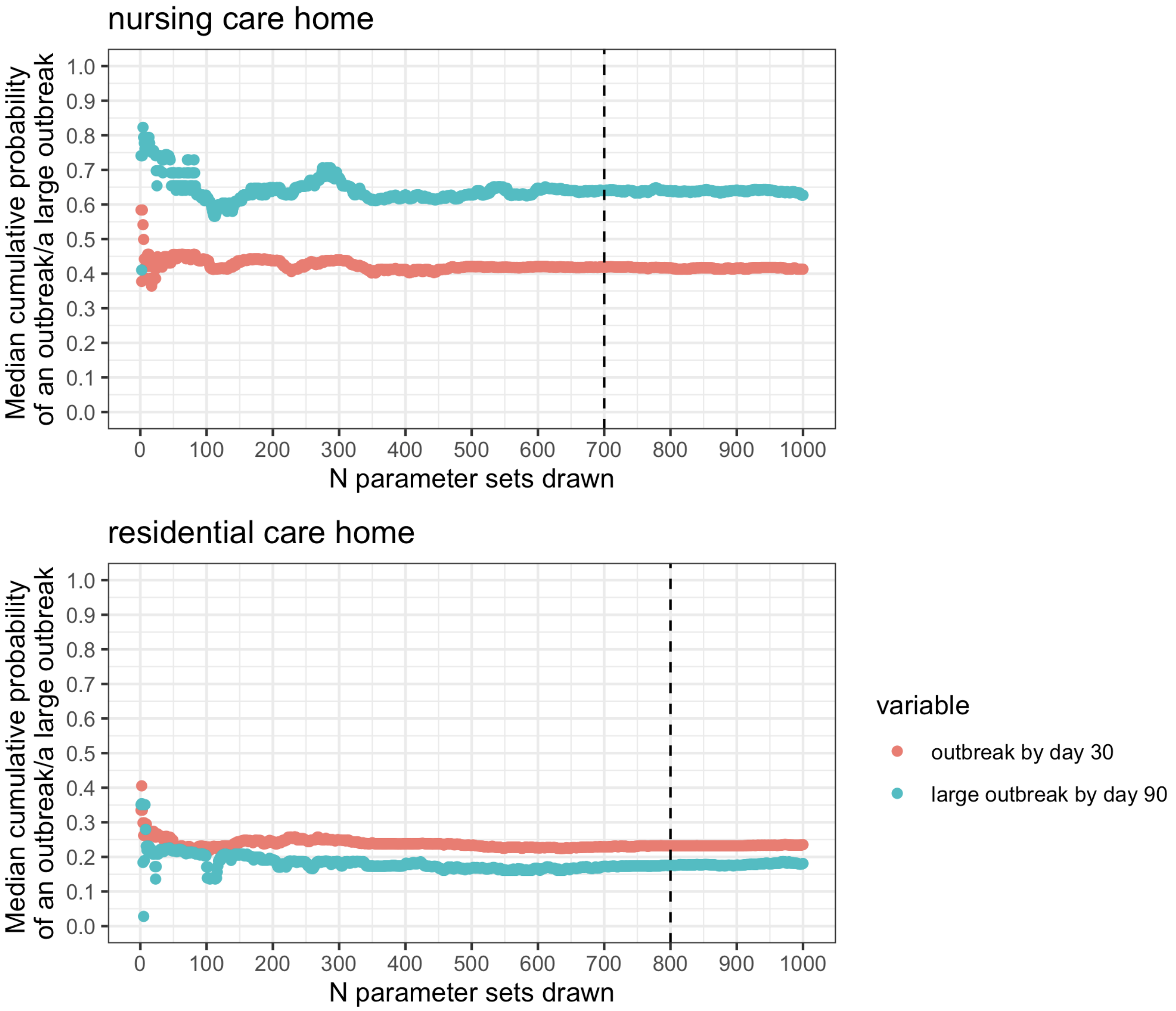


*Figure S4. Cumulative probability of an outbreak by day 30 (red) and a large outbreak by day 90 (blue) in a nursing care home (top plot) and a residential care home (bottom plot) by the number of parameter sets drawn. The dashed line represents the number of parameter sets simulated.*

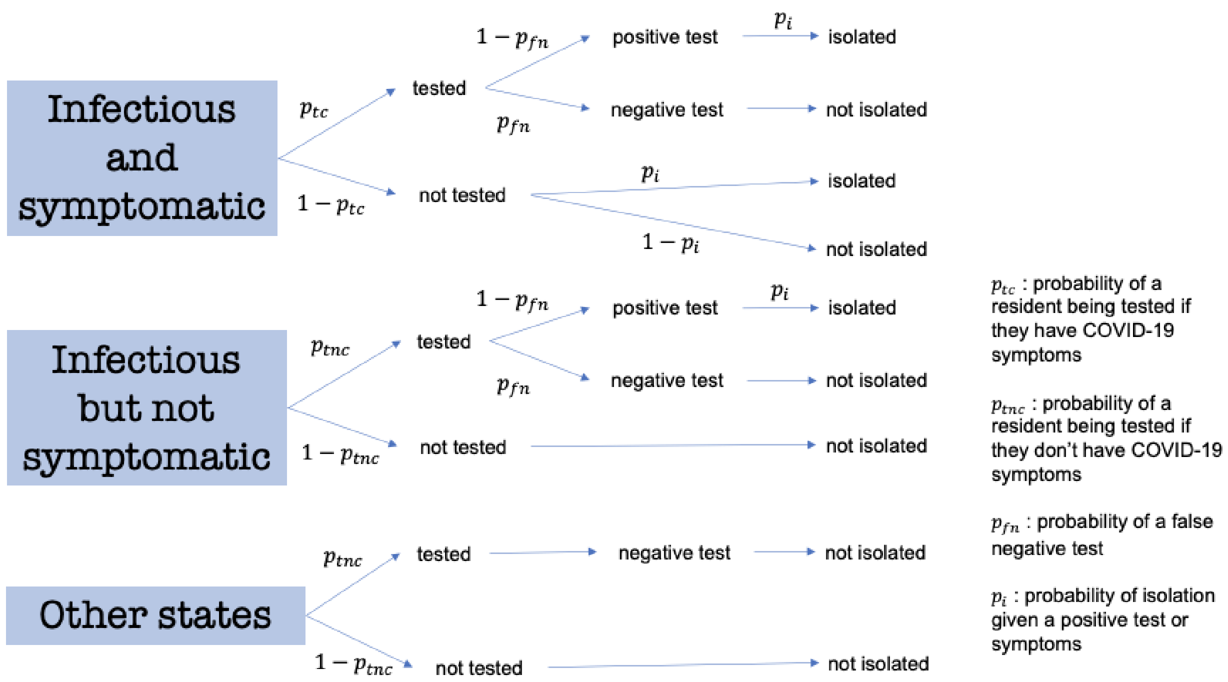


*Figure S5. Testing and isolation pathway for care home residents. Isolation was assumed to be implemented within the care home. The effectiveness of isolation was, on average, 75%.*

.

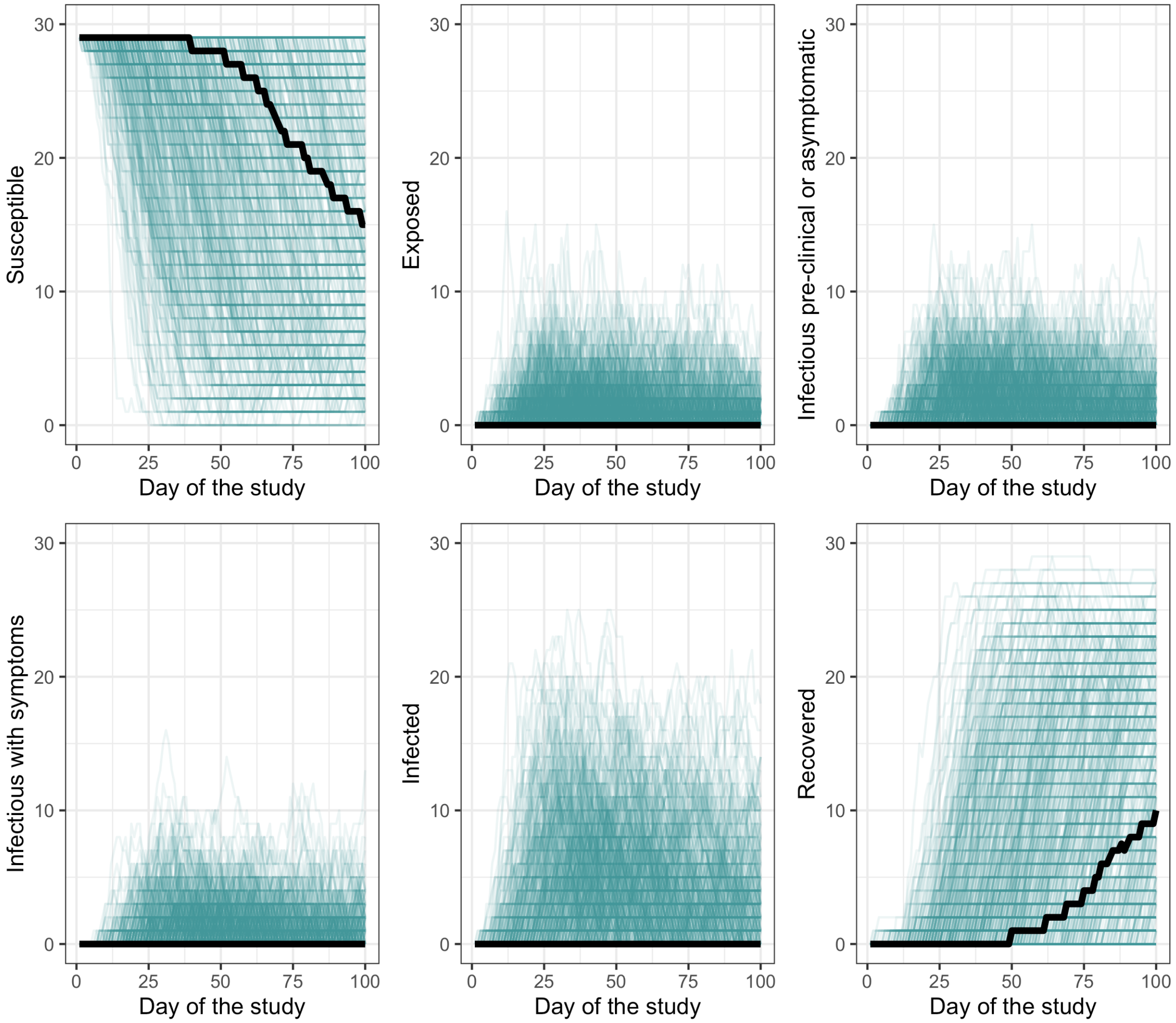


*Figure S6. Care home residents: 1000 model runs by infection state of residents in a residential care home. Infected residents were those either exposed, infectious preclinical, asymptomatic, or symptomatic. The black line represents the median values over time. Day 0 of the study was the day at which the simulations were initialised. Each colour indicates a different run of the model.*

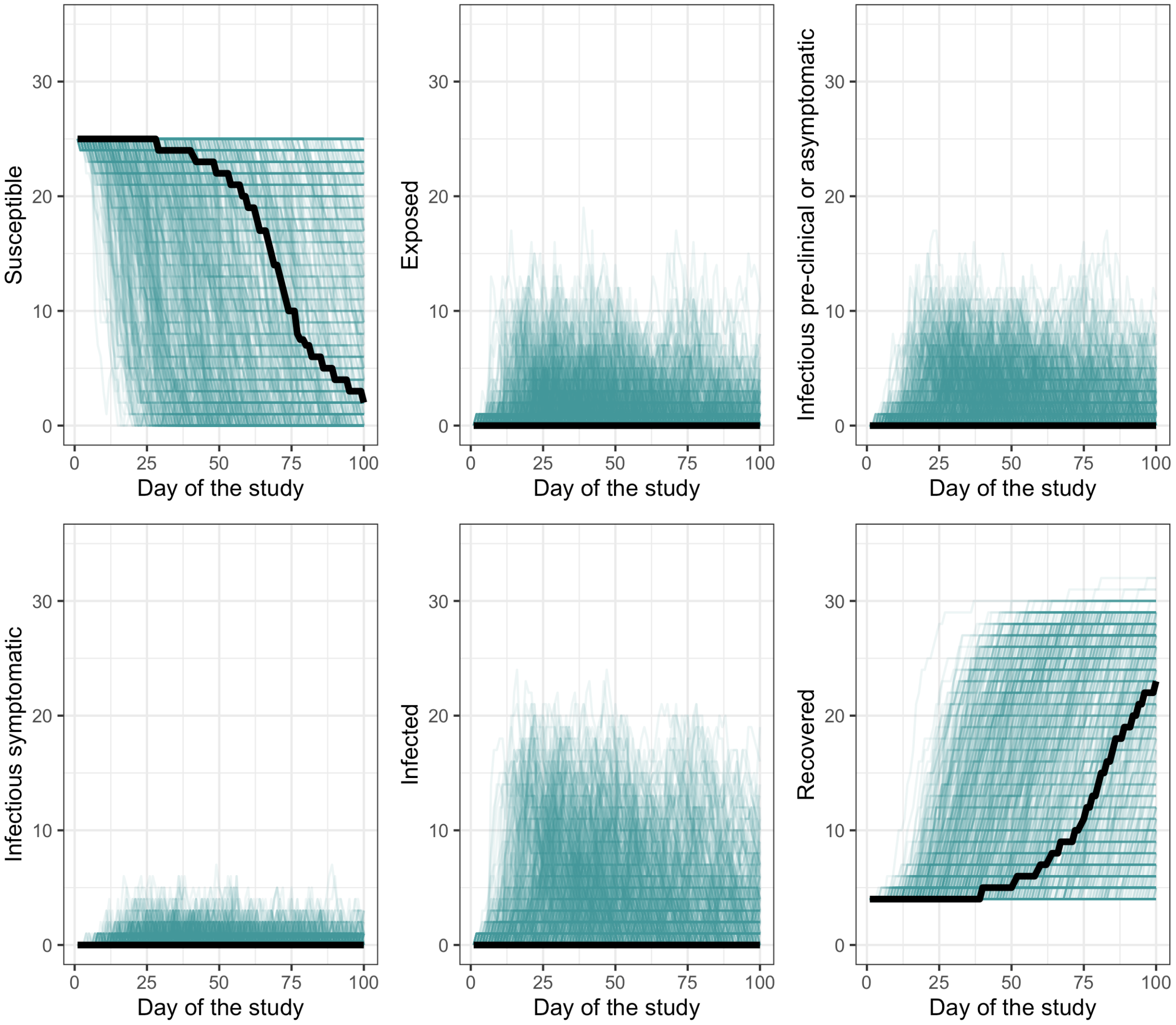


*Figure S7. Care home staff: 1000 model runs by infection state of staff in a residential care home. Infected staff were those either exposed, infectious preclinical, asymptomatic, or symptomatic. The black line represents the median values over time. Day 0 of the study was the day at which the simulations were initialised. Each colour indicates a different run of the model.*

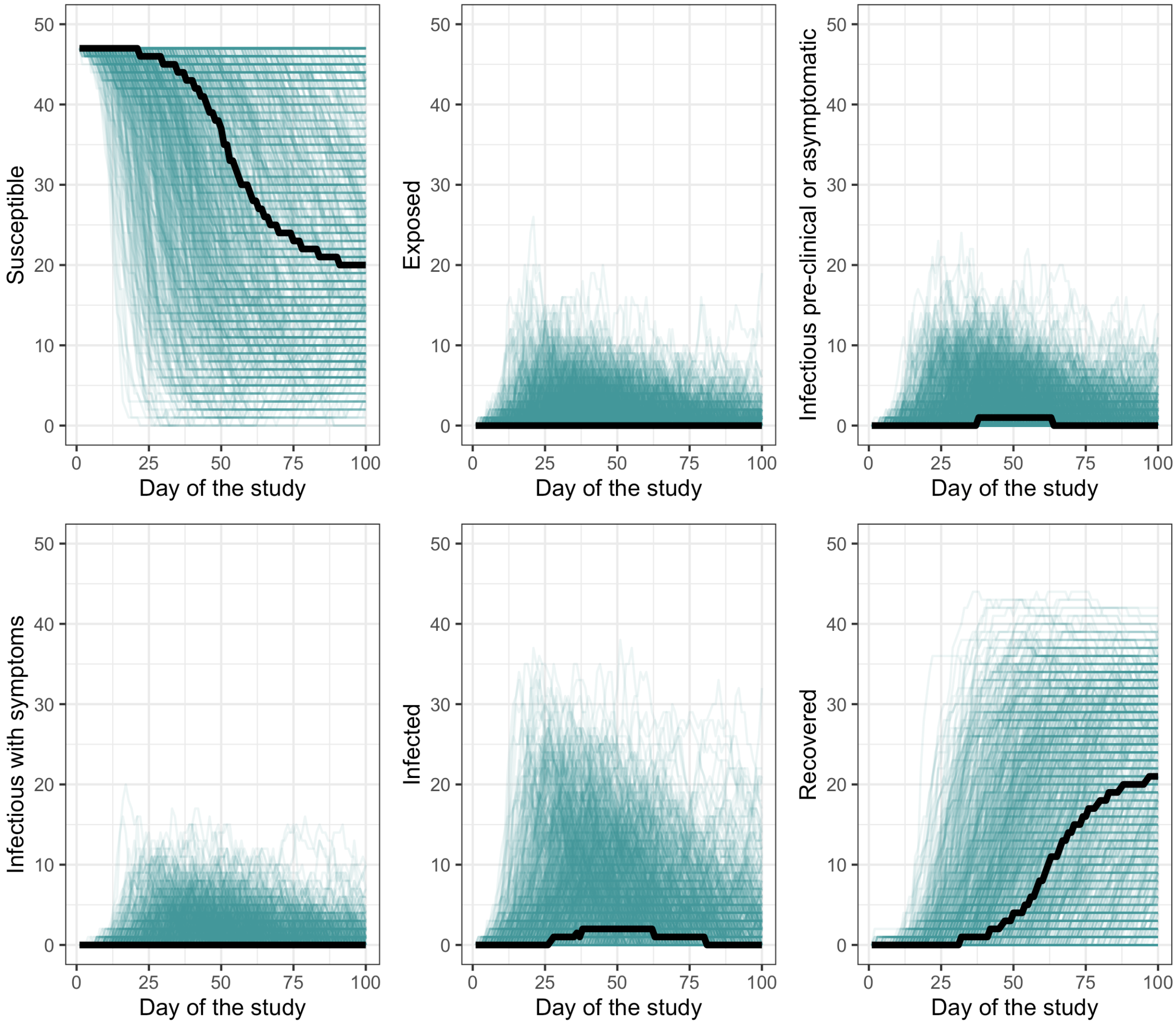


*Figure S8. Care home residents: 1000 model runs by infection state of residents in a nursing care home. Infected residents were those either exposed, infectious preclinical, asymptomatic, or symptomatic. The black line represents the median values over time. Day 0 of the study was the day at which the simulations were initialised. Each colour indicates a different run of the model.*

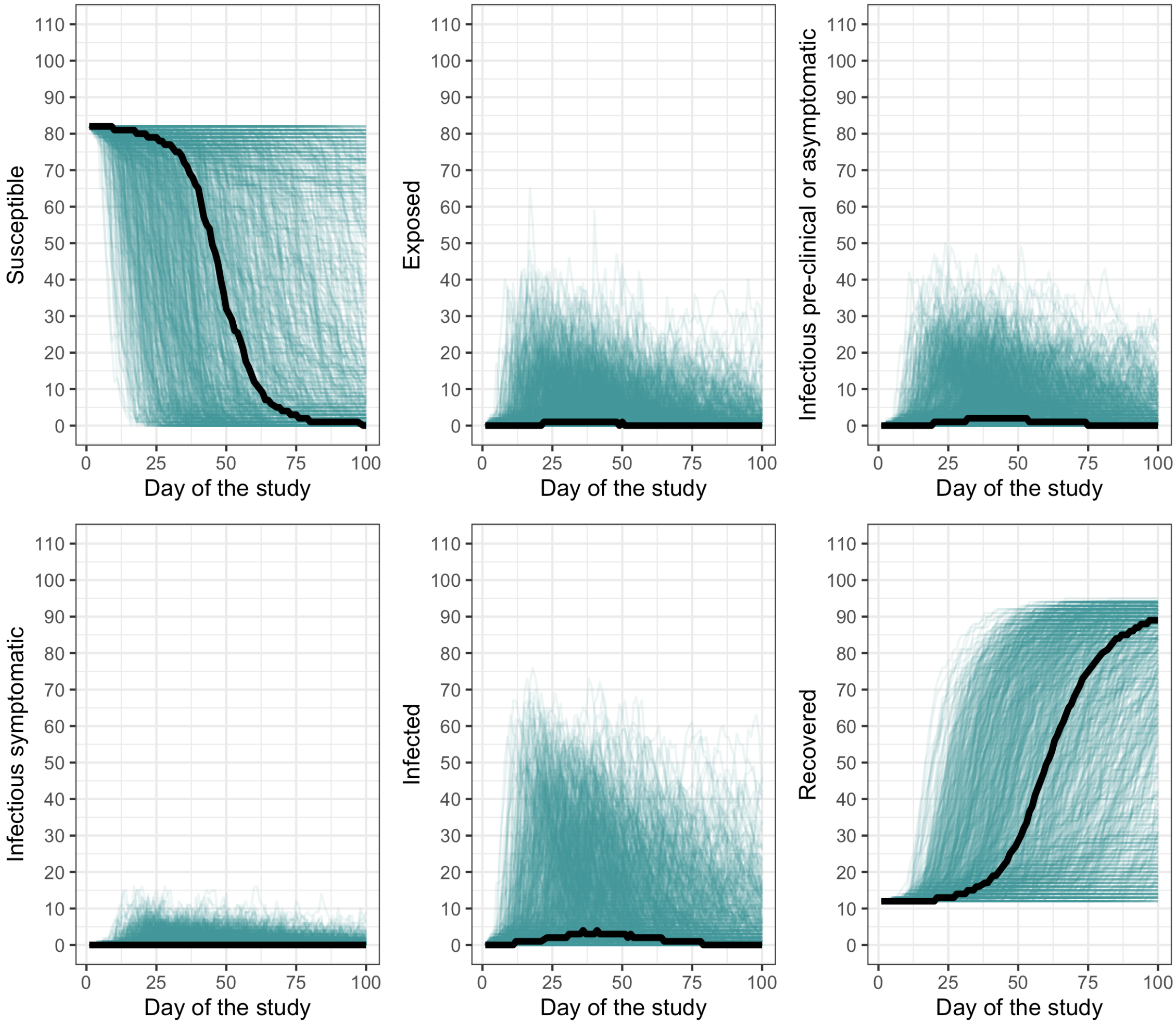


*Figure S9. Care home staff: 1000 model runs by infection state of staff in a nursing care home. Infected staff were those either exposed, infectious preclinical, asymptomatic, or symptomatic. The black line represents the median values over time. Day 0 of the study was the day at which the simulations were initialised. Each colour indicates a different run of the model.*

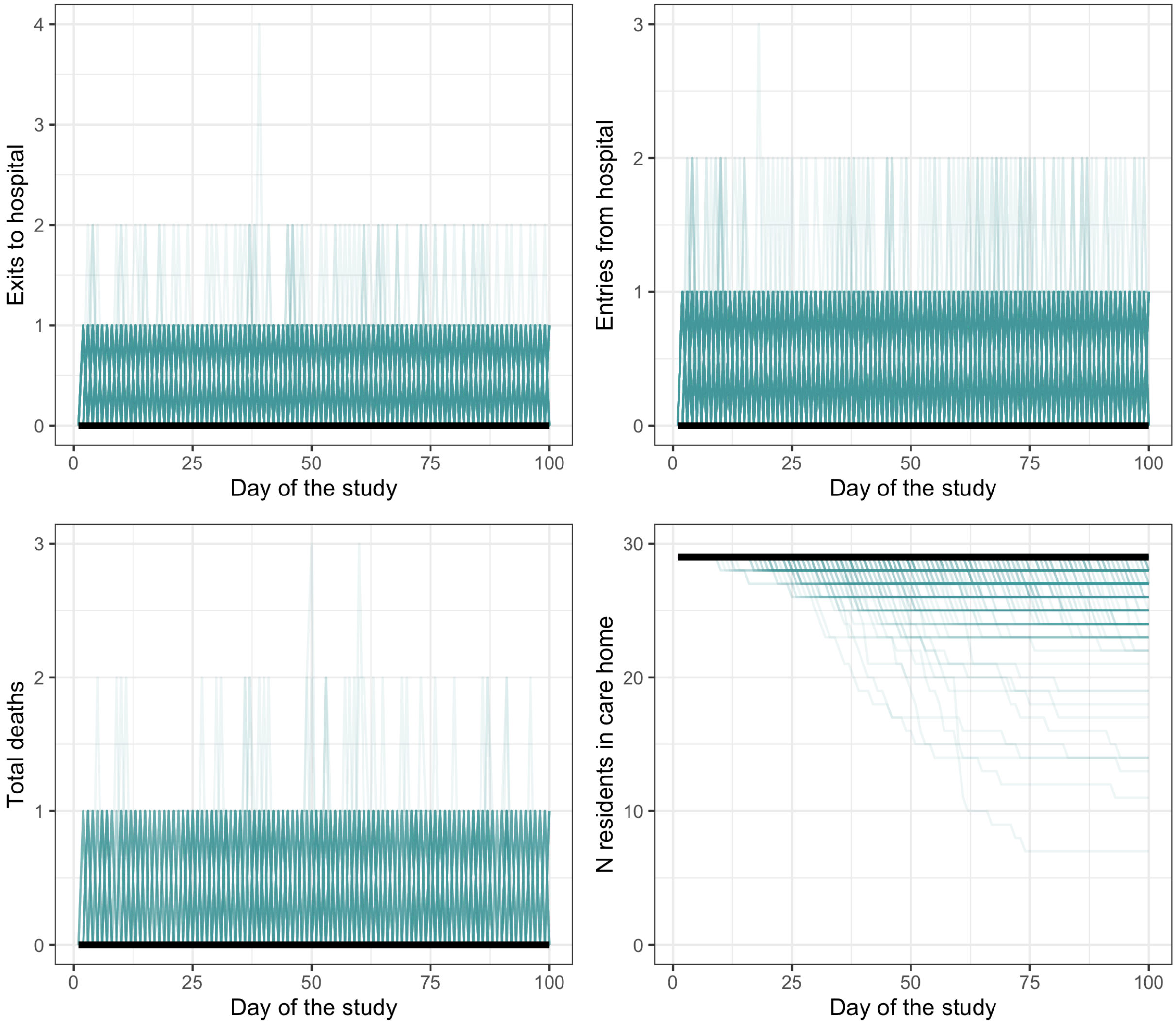


*Figure S10. Resident population in a residential care home for 1000 model runs. The left top panel shows the number of daily exits from the care home to hospital; the right top panel shows the number of daily entries from hospital to the care home, the bottom left panel shows the number of daily deaths in the care home, and the bottom right panel shows the number of residents in the care home each day of the study. The black line represents the median values over time. Day 0 of the study was the day at which the model simulations were initialised. Each colour indicates a different run of the model.*

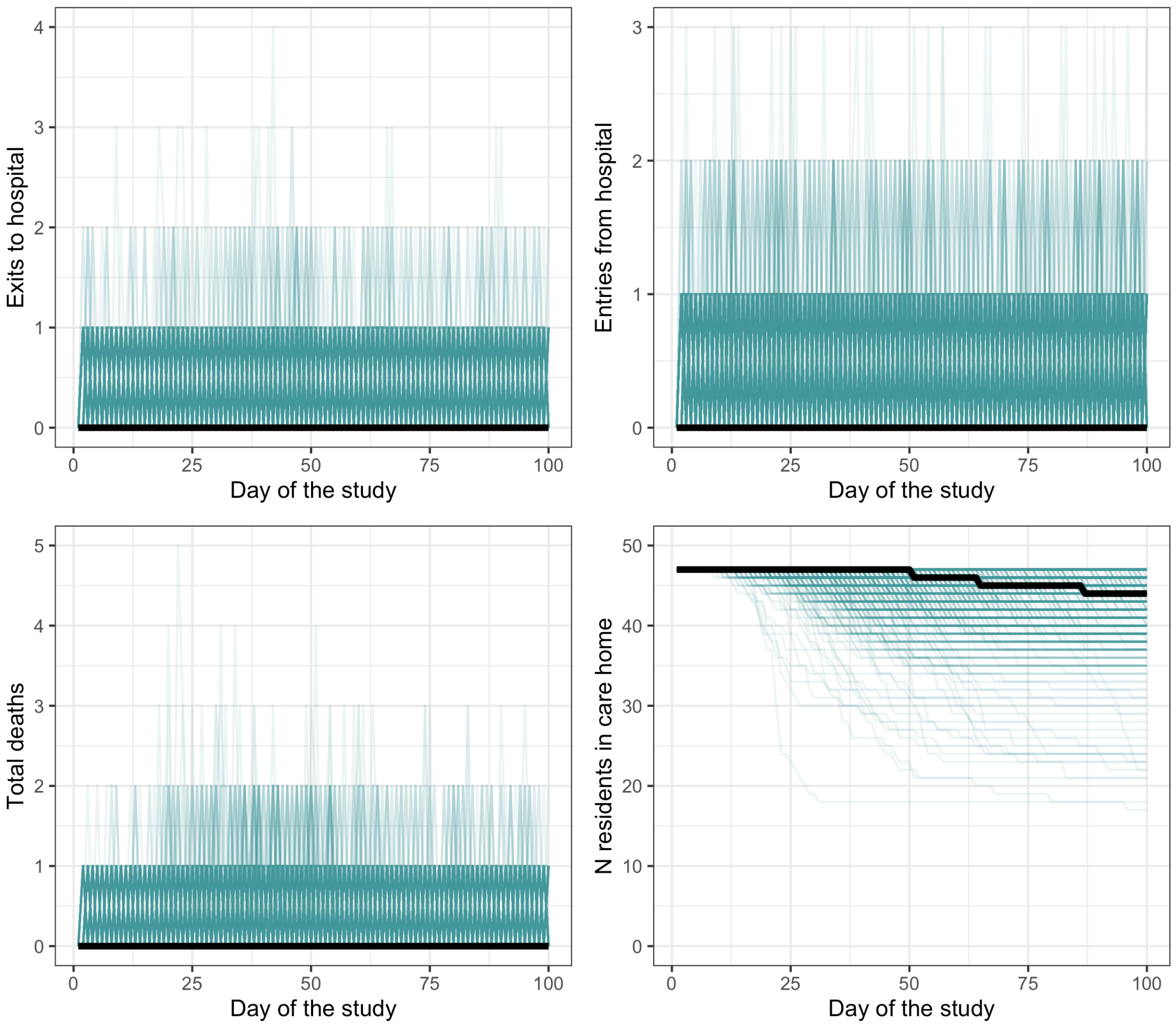


*Figure S11. Resident population in a nursing care home for 1000 model runs. The left top panel shows the number of daily exits from the care home to hospital; the right top panel shows the number of daily entries from hospital to the care home, the bottom left panel shows the number of daily deaths in the care home, and the bottom right panel shows the number of residents in the care home each day of the study. The black line represents the median values over time. Day 0 of the study was the day at which the model simulations were initialised. Each colour indicates a different run of the model.*

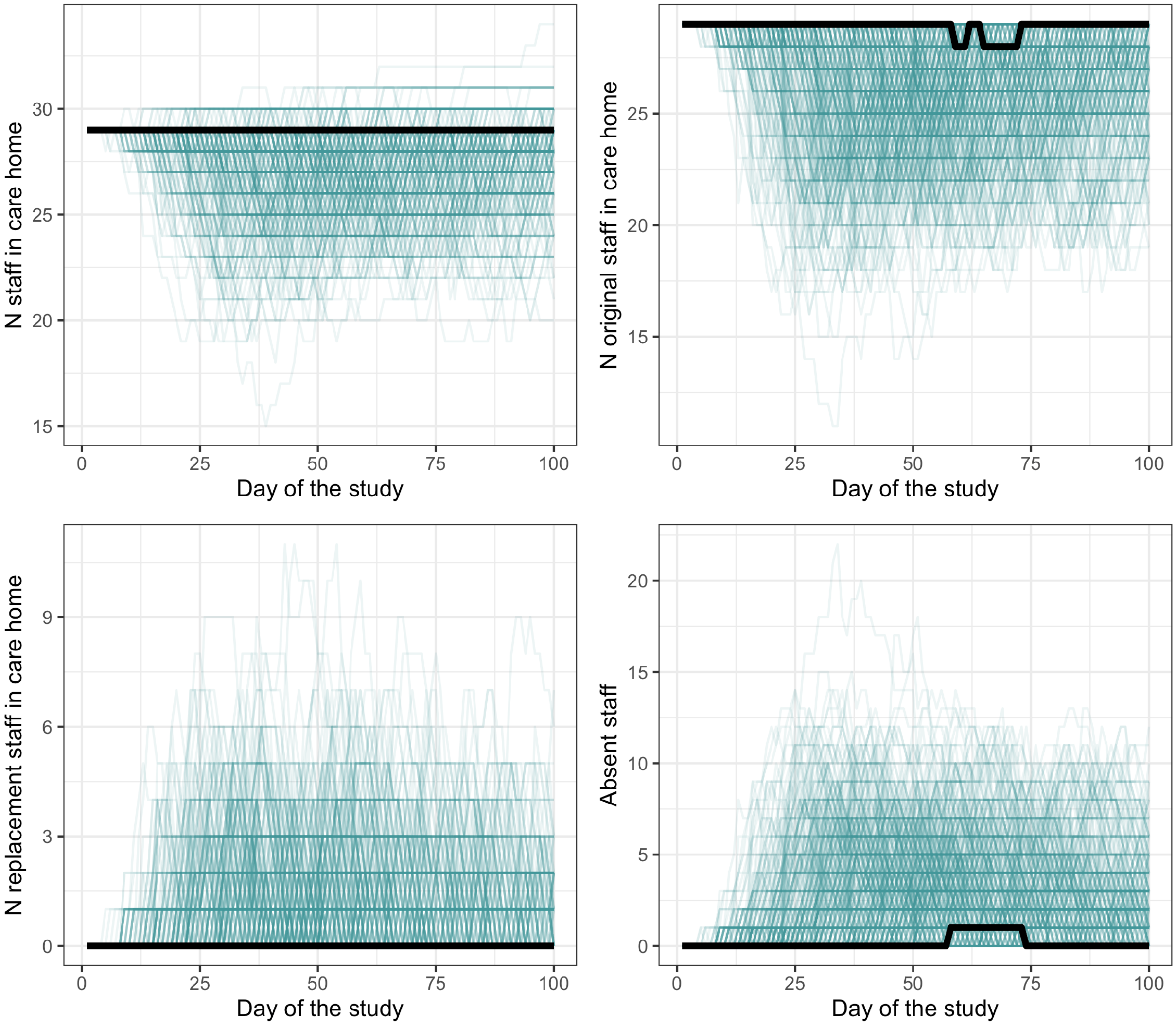


*Figure S12. Staff population numbers in a residential care home for 1000 model runs. The left top panel shows the number of staff in the care home on each day of the study; the right top panel shows the number of original staff in the care home on each day of the study, the bottom left panel shows the number of replacement staff in the care home on each day of the study, and the bottom right panel shows the number of staff absent from the care home on each day of the study. The black line represents the median values over time. Day 0 of the study was the day at which the model simulations were initialised. Each colour indicates a different run of the model.*

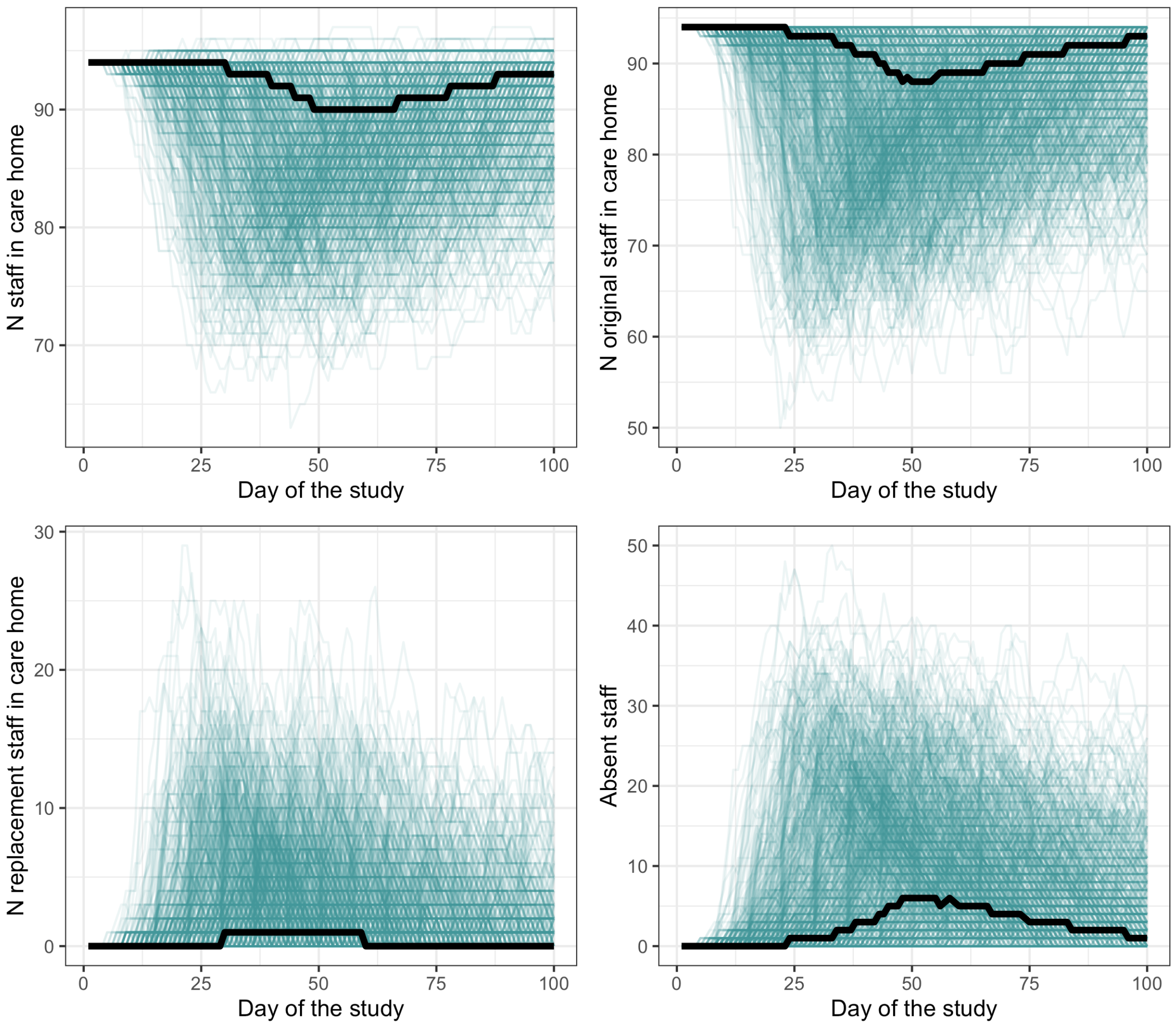


*Figure S13. Staff population numbers in a nursing care home for 1000 model runs. The left top panel shows the number of staff in the care home on each day of the study; the right top panel shows the number of original staff in the care home on each day of the study, the bottom left panel shows the number of replacement staff in the care home on each day of the study, and the bottom right panel shows the number of staff absent from the care home on each day of the study. The black line represents the median values over time. Day 0 of the study was the day at which the model simulations were initialised. Each colour indicates a different run of the model.*

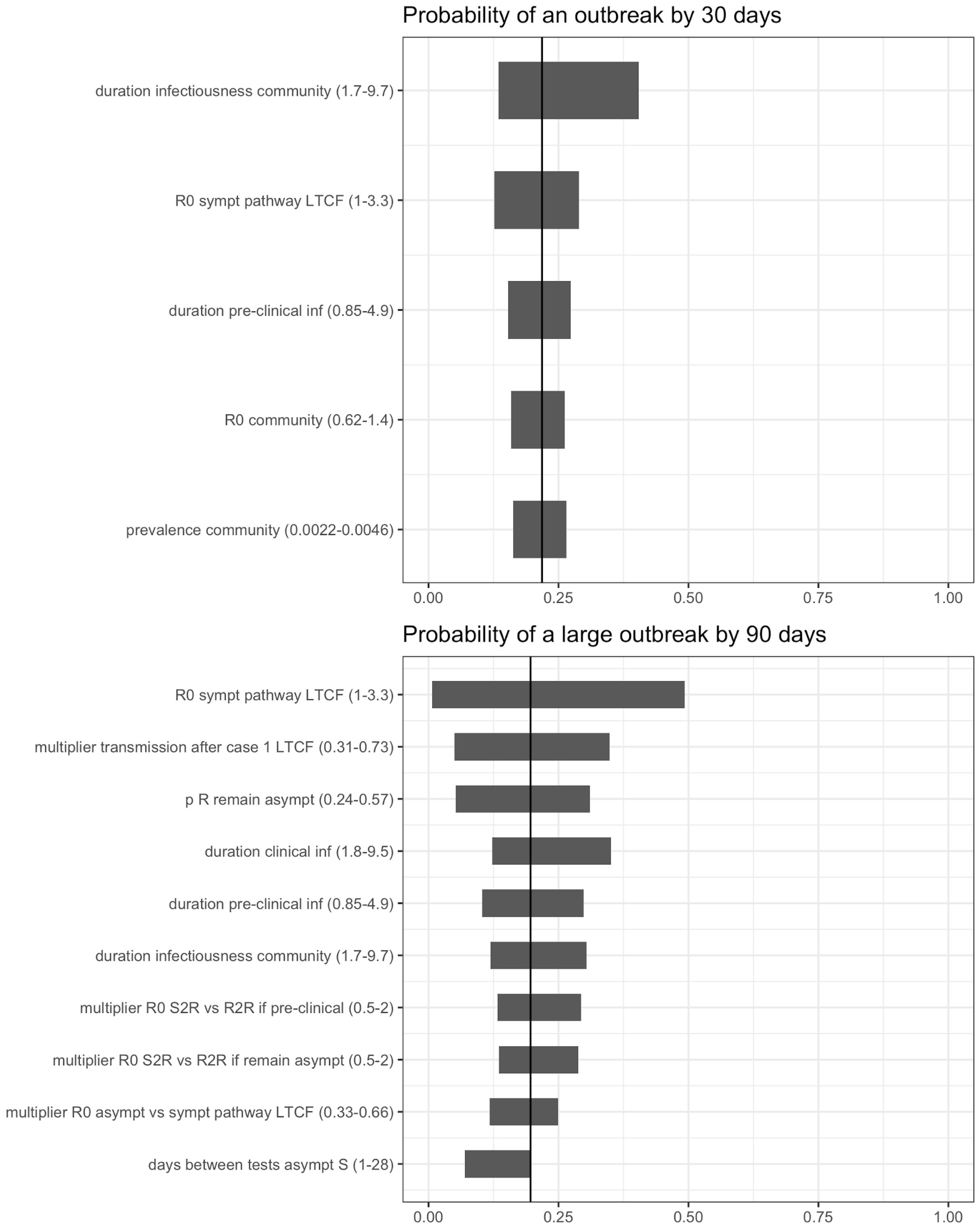


*Figure S14. Sensitivity of model outcomes to model parameters in a residential care home. Only the parameters with the highest impact on model outcomes are shown (>0.1 difference). S=staff; R=resident.*

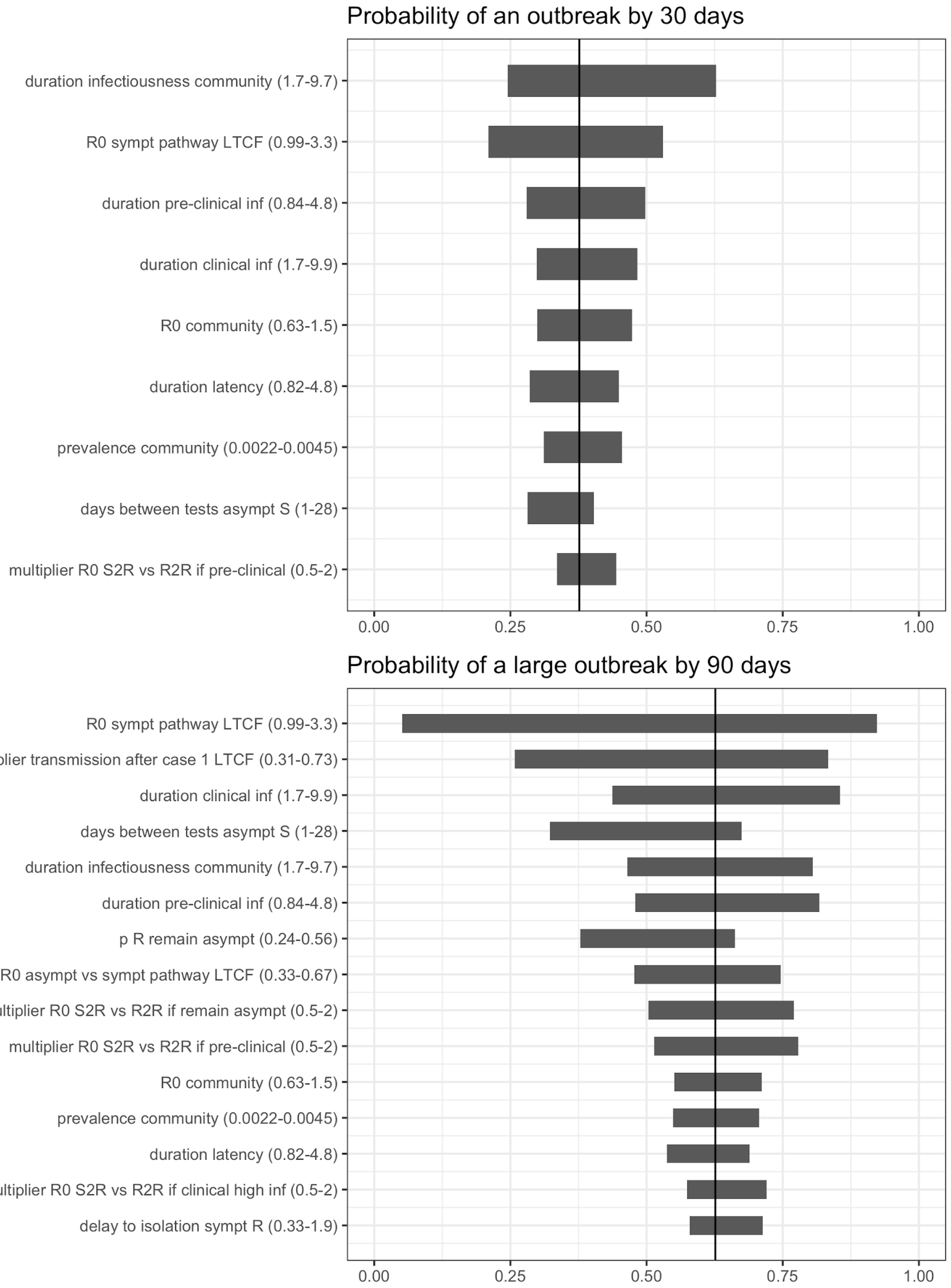


*Figure S15. Sensitivity of model outcomes to model parameters in a nursing care home. Only the parameters with the highest impact on model outcomes are shown (>0.1 difference). S=staff; R=resident.*

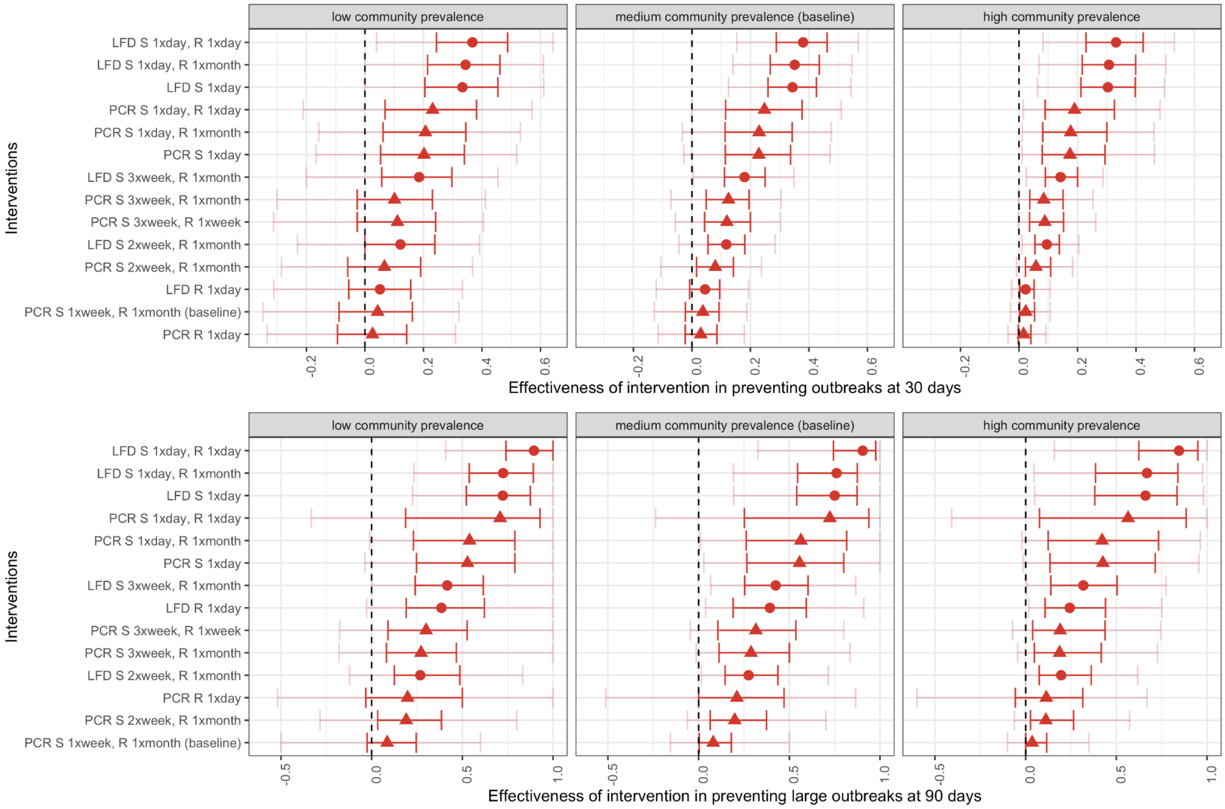


*Figure S16. Effectiveness of testing strategies in preventing outbreaks in residential care homes at 30 days (top panels) and large outbreaks at 90 days (bottom panels) by testing intervention and and under low (left panels), medium (baseline, middle panels) and high (right panels) community prevalence. In red, the 25-75%, in pink, the 5-95%. Testing interventions include PCR testing (triangles) and LFD testing (dots). R stands for resident and S for staff.*

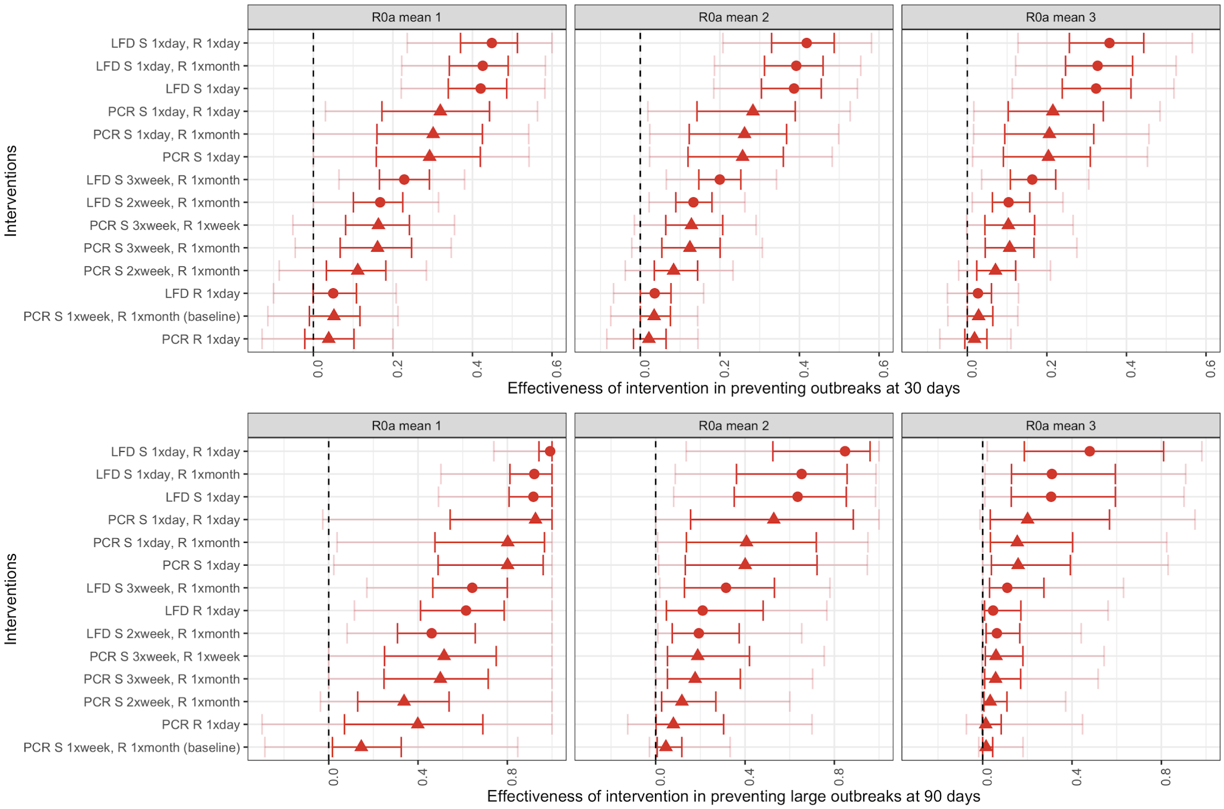


*Figure S17. Effectiveness of testing strategies in preventing outbreaks in nursing care homes at 30 days (top panels) and large outbreaks at 90 days (bottom panels) by testing intervention and and under R0a=1 (left panels), R0a=2 (baseline, middle panels) and R0a=3 (right panels). R0a was the average R0 for individuals who were eventually symptomatic (pathway (a)). In red, the 25-75%, in pink, the 5-95%. Testing interventions include PCR testing (triangles) and LFD testing (dots). R stands for resident and S for staff.*

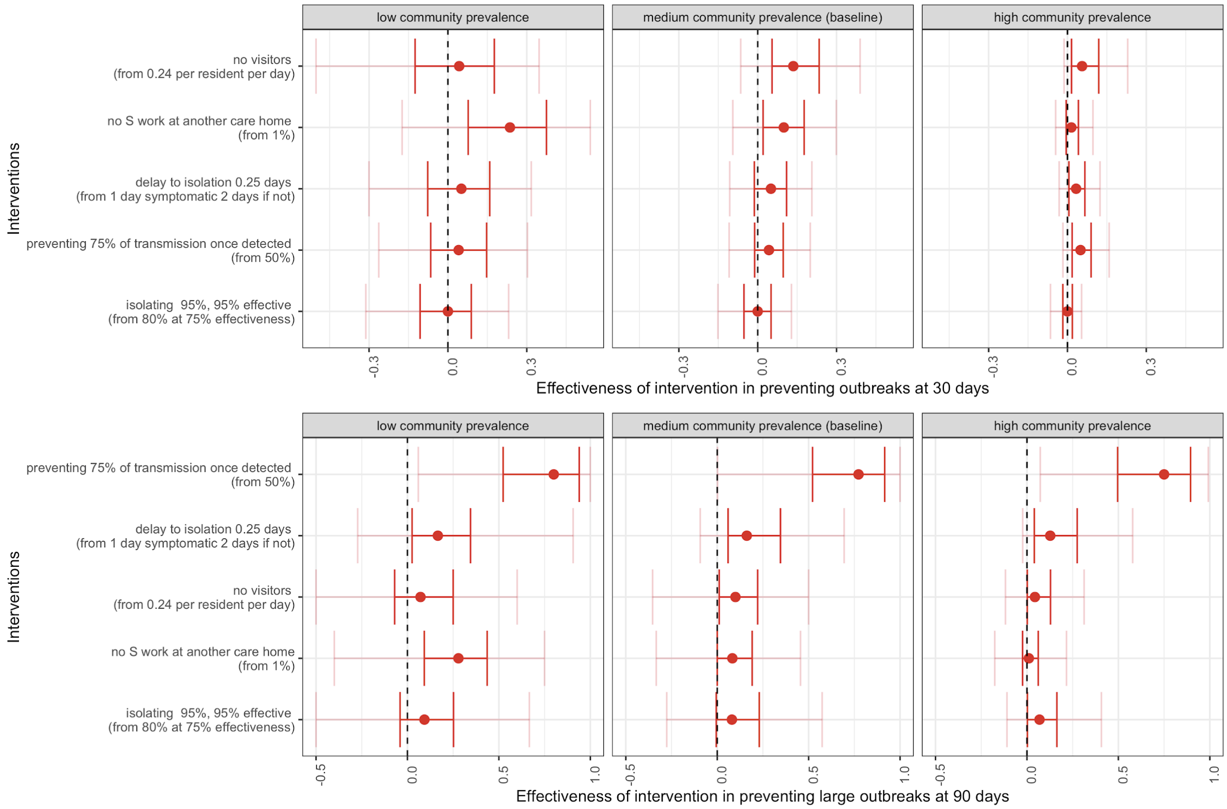


*Figure S18. Effectiveness of interventions in preventing outbreaks in residential care homes at 30 days (top panels) and large outbreaks at 90 days (bottom panels) by low (left panels), medium (baseline, middle panels) and high (right panels) community prevalence. In red, the 25-75%, in pink, the 5-95%. R stands for resident and S for staff.*

#### SUPPLEMENTARY MATERIAL A0

National guidance in England recommends care home providers implement physical distancing, follow shielding guidance, and monitor residents closely for signs of infection (which may be atypical in this population).[18] Residents known to have been exposed to SARS-CoV-2 through contact with a possible or confirmed case of COVID-19 should be isolated or cohorted with other suspected cases where isolation is not possible.[18] Symptomatic residents must be isolated and tested by PCR.[18] Guidance has also been published concerning PPE use for care home staff.[19] Testing upon hospital discharge is required for all care home residents. Since December 2020, upon receipt of a positive test, residents are discharged to “designated settings” (facilities approved by the Care Quality Commission) to isolate for 14 days before returning to their care home.[20] Testing has been provided to symptomatic care home residents and staff since the 8th of April 2020, and from the 6th of July 2020 all staff are recommended to be tested weekly and residents every 28 days, independently of their symptom status.[21] However, delays in the testing pathways have been problematic.[22] On the 23rd of December a policy was introduced to test staff twice a week by LFD (in addition to weekly by PCR).[23] Visiting policies in care homes are decided on an individual basis by care home managers.[24] In the Vivaldi study, which surveyed 9,081 care homes in England from 26 May to 19 June 2020, 97% of care home managers reported their facilities as having been closed to visitors.[25] However, visiting restrictions were eased over the summer of 2020, and lateral flow antigen testing (lateral flow device, LFD) tests are now being used to test all visitors.[21] LFD tests are point of care tests for SARS-CoV-2 that can display results in 15-30 minutes and have been shown to detect a high proportion of the most infectious individuals.[26] However, they have lower sensitivity than laboratory PCR tests for detecting all infections, particularly when carried out by individuals who are insufficiently trained.[27,28]

**SUPPLEMENTARY MATERIAL A1**

***Stochasticity and sensitivity analyses***

Our model was coded in a pomp framework in R using C snippets[29] with a time step of one day. The code is available here (public once published):

https://github.com/rmjlros/COVID19_care_home_NPIs.

The model was initialised with all residents being susceptible. The initial proportion of staff in each compartment was informed by results from a dynamic mathematical model of SARS-CoV-2 transmission calibrated to community data[5].

The stochastic draw determining the number of individuals moving through each transition at each timestep was simulated through Euler-multinomial transitions using the "reulermultinom" function of the pomp package in R.[29] Other entries and exits into the care home were modelled using random Poisson processes using the "rpois" function.

We randomly drew 600 parameter sets and, for each, ran 700 simulations for nursing care homes homes and 800 simulations for residential care homes. The number of parameter sets and simulations per parameter set were determined by examining the point at which the model outputs converged (see Figures S3 and S4).

Univariate sensitivity analyses were carried out to identify the parameters that model outcomes were most sensitive to. We fixed parameter values at the 5th and 95th percentile of their distribution (if the parameter was drawn from a distribution), or selected plausible ranges if they were not, and outputted the probability of an outbreak and of a large outbreak for each parameter value selected.

***Natural history of SARS-CoV-2 infection and transmission process***

Susceptible residents could become exposed to SARS-CoV-2 through contact with infectious residents and staff in the care home, and through contact with infectious visitors (see below for details). Susceptible staff could become exposed through contact with infectious residents and staff, and through their work in other care homes, and outside of care homes in the community. Exposed residents and staff progressed to either pathway (a), exhibiting preclinical (asymptomatic) infection followed by clinical (symptomatic) infection, or pathway (b), exhibiting asymptomatic infection only.

Infected residents remained in the care home unless hospitalized (described below). Infected staff were removed from the care home if their infection was detected either through testing (described below) or as a result of their symptoms. Staff with asymptomatic infection continued to work until they received a positive test, and otherwise remained infectious within the care home until their recovery. Absent staff were assumed to be absent for a mean duration of 14 days (95% 10.5-17.8) before returning to the care home in a recovered state. Transmission rates were, on average, halved (95% 31-73%) whilst a care home had a detected outbreak (i.e. whilst one or more residents were isolated or one or more members of staff were absent), in order to reflect a step up in IPC measures due to increased awareness of SARS-CoV-2 infection in the care home but a reflection of the difficulty in IPC in this care setting (see $m\beta_d$ in Table S1).

A mean of 80% (95% 72-88%) of absent care home staff were assumed to be replaced by a second pool of staff. The remaining absent staff were not replaced and only resumed work after a mean of 14 days, during which there was a staff vacancy in the care home. When replacement staff became absent they did not return to the care home after their isolation period was completed (since the replacement was temporary). Replacement staff exited the care home at the same rate that the original staff members recovered and re-entered the care home. Original and replacement staff were considered to be the same in relation to force of infection calculations, except replacement staff were more likely to work at more than one care home and hence have a higher probability of being infected (details below).

***Residents***

Susceptible residents could become exposed to SARS-CoV-2 through contact with infectious residents ($\lambda_{r2r}$) and staff ($\lambda_{s2r}$) in the care home, and through contact with infectious visitors ($\lambda_{v2r}$) (see Supplementary material A1 for details). Susceptible staff could become exposed through contact with infectious residents($\lambda_{r2s}$) and staff ($\lambda_{s2s}$), and through their work in other care homes ($\lambda_{o2s}$), and outside of care homes in the community ($\lambda_{c2s}$).

Exposed residents progressed to either pathway (a), exhibiting preclinical (asymptomatic) infection followed by clinical (symptomatic) infection, or pathway (b), exhibiting asymptomatic infection only. Exposed residents progressed to pathways (a) and (b) at proportions $1-p_a$ and $p_a$, respectively, after a latent period $1/\varepsilon$. Infected pre-clinical residents progressed to clinical infection at rate $\tau$ (symptom onset). Clinical infection was further subdivided into two periods: early clinical infection with high infectiousness followed by late clinical infection with low infectiousness, each having the same duration ($1/\omega$ and $1/\varphi$). This enabled better characterisation of viral load peaking at symptom onset and decreasing rapidly thereafter [2,3]. Residents recovered from infection at a recovery rate $\varphi$ and were considered immune from future SARS-CoV-2 infection for the duration of the simulation. The duration of asymptomatic infection $1/\varphi_a$ (pathway (b)) was assumed to equal the duration of the combined preclinical and clinical duration $1/\tau + 1/\omega + 1/\varphi$(pathway (a)).

Transmissibility of SARS-CoV-2 was assumed to vary over the different stages of pathway (a) (pre-clinical, early clinical, and late clinical stages). Conversely, for pathway (b), transmissibility was assumed to remain at the same rate over the full duration of infectiousness.

The overall force of infection between residents $\lambda_{r2r}$ was calculated as follows (simplified, see full equation below):

$$\lambda_{r2r} = \beta_{pc}\frac{Ipc}{N_r} + \beta_{ch}\frac{Ich}{N_r} + \beta_{cl}\frac{Icl}{N_r} + \beta_{a}\frac{Ia}{N_r}$$

where $Ipc$ represents the number of residents in the infectious pre-clinical compartment, $Ich$ is the number of residents in the infectious clinical with high infectiousness compartment, $Icl$ is the number of residents in the infectious clinical with low infectiousness compartment, $Ia$ is the number of residents in the infectious asymptomatic compartment, and $N_r$ represents the total number of residents present in the care home. Among individuals in pathway (a) ie. those who eventually present with symptoms (individuals go through pre-clinical, early clinical, and late clinical stages), the transmission rate was subdivided into $\beta_{pc}$ for preclinical SARS-CoV-2 infection, $\beta_{ch}$ for early clinical infection and $\beta_{cl}$ for later clinical infection. For pathway (a), we assumed a total reproduction number $R0_a$ (R0 for pathway (a)) over the full infectious period. The total duration of infectiousness was subdivided into a preclinical infection period, a period of clinical symptomatic infection with high levels of infectiousness and a period of clinical symptomatic infection with low levels of infectiousness. Assuming that on average that a proportion $p_{transmission\_pc\_vs\_c}$ of transmission occurs during preclinical infection compared to clinical infection and that a proportion $p_{transmission\_cl\_vs\_ch}$ of transmission occurs in the later stage of clinical infection compared to the early stage of clinical infection, we find:

$$\beta_{pc} = \frac{R0_a \times p_{transmission\_pc\_vs\_c}}{d\_preclinical\_infection};$$

$$\beta_{ch} = \frac{R0_a \times (1 - p_{transmission\_pc\_vs\_c}) \times (1 - p_{transmission\_cl\_vs\_ch})}{d\_clinical\_infection\_h};$$

$$\beta_{cl} = \frac{R0_a \times (1 - p_{transmission\_pc\_vs\_c}) \times p_{transmission\_cl\_vs\_ch}}{d\_clinical\_infection\_l}.$$

A separate transmission rate was estimated for pathway (b), where residents remain asymptomatic during the full duration of their infection. This transmission rate was derived assuming $R0_b$ over the full asymptomatic infectious period:

$$\beta_a = \frac{R0_b}{d\_total\_infectiousness},$$

where $R0_b = R0_a \times m_{R0_b}$ and $0 < m_{R0_b} < 1$.

The full equation for the force of infection between residents $\lambda_{r2r}$ is as follows:

$$\lambda_{r2r} = \beta_{pc} \frac{Ipc + Ipcpi + (Ipci \times m_i)}{N_r} + \beta_{ch} \frac{Ich + Ichpi + ((Ichi) \times m_i)}{N_r} + \beta_{cl} \frac{Icl + Iclpi + (Icli \times m_i)}{N_r} + \beta_a \frac{Ia + Iapi + (Iai \times m_i)}{N_r},$$

where $\beta_{pc}$ was the transmission rate for those infectious pre-clinical, $Ipc$ was the number of residents infectious pre-clinical that would not be isolated, $Ipcpi$ was the number of residents infectious pre-clinical that would be but were not yet isolated, $Ipci$ was the number of residents infectious pre-clinical isolated, $m_i$ was the relative infectiousness of residents isolated compared to those that were not, $N_r$ was the number of residents in the care home, $\beta_{ch}$ was the transmission rate for those highly infectious presenting with clinical symptoms, $Ich$ was the number of residents highly infectious presenting with clinical symptoms that would not be isolated, $Ichpi$ was the of number residents highly infectious presenting with clinical symptoms that would be but were not yet isolated, $Ichi$ was the number of residents highly infectious presenting with clinical symptoms that were isolated, $\beta_{cl}$ was the transmission rate for those less infectious presenting with clinical symptoms $Icl$ was the number of residents less infectious presenting with clinical symptoms that would not be isolated, $Iclpi$ was the number of residents less infectious presenting with clinical symptoms that would be but were not yet isolated, $Icli$ was the number of residents less infectious presenting with clinical symptoms isolated, $\beta_a$ was the transmission rate for those asymptomatic, $Ia$ was the number of asymptomatic residents that would not be isolated, $Iapi$ was the number of asymptomatic residents that would be but were not yet isolated, $Iai$ was the number of asymptomatic residents isolated.

In the baseline scenario, residents could become exposed to infection through contact with other (infectious) residents or staff. Therefore, the total rate of exposure of residents $\lambda_R$ was defined as:

$$\lambda_R = \lambda_{r2r} + \lambda_{s2r} + \lambda_{v2r},$$

where $\lambda_{r2r}$ describes the force of infection from residents to residents (described above), $\lambda_{v2r}$ described the force of infection from visitors to residents (described below), and $\lambda_{s2r}$ describes the force of infection from staff to residents, calculated as follows (simplified, see full equation below):

$$\lambda_{s2r} = \beta_{pc}\frac{Ipcs}{N_s} + \beta_{ch}\frac{Ichpis}{N_s} + \beta_a\frac{Ias}{N_s},$$

where the transmission rates are calculated as described above for $\lambda_{r2r}$ and $N_s$ describes the total number of staff in the care home, including replacement staff. $Ipcs$ were staff in the infectious pre-clinical compartment, $Ias$ were staff in the infectious asymptomatic compartment, and $Ichpis$ describes symptomatic and highly infectious staff who were present in the care home before becoming absent after a delay $1/\gamma_c$.

The full equation for the force of infection from staff to residents $\lambda_{s2r}$ is as follows:

$$\lambda_{s2r} = \beta_{pc}\frac{Ipcs+Ipcpis+Ipcs2+Ipcpis2}{N_s} + \beta_{ch}\frac{Ichpis+Ichpis2}{N_s} + \beta_a\frac{Ias+Iapis+Ias2+Iapis2}{N_s},$$

where $\beta_{pc}$ was the transmission rate for those infectious pre-clinical, $Ipcs$ was the number of original staff infectious pre-clinical that would not be absent, $Ipcpis$ was the number of original staff infectious pre-clinical that would be but were not yet absent, $Ipcs2$ was the number of replacement staff infectious pre-clinical that would not be absent, $Ipcpis2$was the number of replacement staff infectious pre-clinical that would be but were not yet absent, $N_s$was the number of staff in the care home, $\beta_{ch}$was the transmission rate for those highly infectious presenting with clinical symptoms, $Ichpis$ was the of number original staff highly infectious presenting with clinical symptoms that would be but were not yet absent, $Ichpis2$ was the of number replacement staff highly infectious presenting with clinical symptoms that would be but were not yet absent,$\beta_a$was the transmission rate for those asymptomatic, $Ias$was the number of asymptomatic original staff that would not be absent, $Iapis$ was the number of asymptomatic original staff that would be but were not yet absent, $Ias2$was the number of asymptomatic replacement staff that would not be absent, $Iapis2$ was the number of asymptomatic replacement staff that would be but were not yet absent.

***Staff***

Staff could become infected through contact with residents, through contact with other staff, through their work in other care homes, or outside of care homes (in the community). Therefore, the total rate of exposure of staff $\lambda_S$ was defined as:

$$\lambda_S=\lambda_{r2s}+\lambda_{s2s} + \lambda_{c2s} + \lambda_{o2s},$$

where$\lambda_{r2s}$represents the force of infection from residents to staff, which was assumed to be equal to the force of infection from residents to residents $\lambda_{r2r}$(described above); $\lambda_{s2s}$describes the force of infection from staff to staff, assumed to be equal to the force of infection from staff to residents $\lambda_{s2r}$(described above); $\lambda_{c2s}$ was the force of infection from the community to staff and $\lambda_{o2s}$describes the force of infection to staff from their work in other care homes.$\lambda_{c2s}$ was calculated as follows:

$$\lambda_{c2s} = \frac{R_C}{d\_total\_infectiousness\_C} \times \ p_{C_{infectious}},$$

where $R_C$ was the reproduction number in the community, assumed in the baseline scenario (medium community prevalence) to follow a Gamma distribution of mean 1, and $d\_total\_infectiousness\_C$ was the duration of infectiousness in the community, assumed to follow a Gamma distribution of mean of 5 days[4]. The probability of an individual in the community being infectious ($p_{C_{infectious}}$) was derived from a dynamic mathematical model of infection calibrated to community data[4] and the same distribution was throughout our simulations for that specific scenario (varied for the low and high community prevalence scenarios).

On average 1% of original staff were assumed to work in another care home (pers. comm. lead of care home survey Thames Valley, Dr Conall Watson, June 2020). On average, we assumed 20% of replacement care home staff worked at another care home. The force of infection to staff working in another care home, $\lambda_{o2s}$, was calculated as:

$$\lambda_{o2s} = (\lambda_{or2s} + \lambda_{os2s}) = [\beta_{pathway\ a} \times p_{res\ infectious\ pathway\ a} + \beta_{pathway\ b} \times p_{res\ infectious\ pathway\ b}] + \dots$$
$$\dots + [\beta_{pathway\ a} \times p_{staff\ infectious\ pathway\ a} + \beta_{pathway\ b} \times p_{staff\ infectious\ pathway\ b}],$$

where $\beta_{pathway\ a}$ and $\beta_{pathway\ b}$ were defined as:

$$\beta_{pathway\ a} = \frac{R0_a}{d\_total\_infectiousness},\ \beta_{pathway\ b} = \frac{R0_b}{d\_total\_infectiousness},$$

and $p_{res\ infectious\ pathway\ a}$ and $p_{res\ infectious\ pathway\ b}$ were the probability of a resident being infectious in pathway (a) and pathway (b), respectively, and $p_{staff\ infectious\ pathway\ a}$ and $p_{staff\ infectious\ pathway\ b}$ were the probability of a staff member being infectious in pathway (a) and pathway (b), respectively.

We make the following assumptions regarding the probability of residents and staff being on different infectious pathways:

$$p_{res\ infectious\ pathway\ a} = p_{care\ home\ outbreak} \times p_{res\ inf} \times (1 - p_a),$$

$$p_{res\ infectious\ pathway\ b} = p_{care\ home\ outbreak} \times p_{res\ inf} \times p_a,$$

$$p_{staff\ infectious\ pathway\ a} = p_{staff\ infectious\ pathway\ b} = p_{care\ home\ outbreak} \times p_{staff\ inf} \times p_{as},$$

where $p_{care\ home\ outbreak}$ was the probability of another care home has an outbreak on a given day, $p_{res\ inf}$ is the probability that a resident in a care home with an outbreak is infectious, $p_{staff\ inf}$ is the probability that a staff member in a care home with an outbreak is infectious, $p_a$ is the probability that a resident follows pathway (a), and $p_{as}$ is the probability that a staff member follows pathway (a).

In the baseline scenario (medium community prevalence), we assumed that the mean probability of a care home having an outbreak on any given day was 25% (corresponding to ~4,000 out of ~15,000 care homes having an outbreak), and the mean probability of a resident (or staff member) being infectious within a care home with an outbreak was 10%, corresponding approximately to 3 residents or staff in a care home being infectious in an average sized residential care home. As denoted above, 50% of staff and 40% of residents were assumed to never present with symptoms (i.e. to be in pathway (b)).

The proportion of replacement staff entering each infection state compartment was assumed to be the same as the proportion of the general community in each disease compartment, and informed by a dynamic mathematical model of infection calibrated to community data[5].

***Visitors***

In the scenarios including visitors, residents could additionally become exposed through contact with visitors:

$$\lambda_{v2r} = \boldsymbol{\beta}_a \frac{Nvisitors}{N_r}$$

The transmission rate for asymptomatic infection ($\boldsymbol{\beta}_a$) was also used for visitors, thus we assumed that any infected visitors were asymptomatic if permitted entry into the care home (for simplicity, we did not allow visitors to be in the preclinical stage of COVID-19 infection). The number of infected visitors per day in the care home, $Nvisitors,$ was given by:

$$Nvisitors = n_V \times Nr \times p_{v_{infectious}},$$

where $n_V$ was the expected number of visitors per resident per day, and $p_{v_{infectious}}$ represents the probability of a visitor being infectious, which we assumed was the same as the probability of being infectious without symptoms in the community overall, and was parameterised using a SARS-CoV-2 transmission model calibrated to community data[30]. Staff were assumed not to become exposed through contact with visitors.

#### SUPPLEMENTARY MATERIAL A2

##### *Mortality and hospitalisation dynamics*

Residents were assumed to exit the care home either due to hospitalisation (either for COVID-19 or other reasons) or death. Residents entered the facility exclusively from hospital (it was assumed that during the COVID-19 pandemic, community admissions of residents to care homes were rare). We also assumed that the proportion of residents leaving the care home to return to the community or transfer to another care home was negligible. Residents without symptomatic COVID-19 (i.e. those in the $S$, $E$, $I_{pc}$, $I_a$ and $R$ compartments) were assumed to be hospitalised at background rate $\kappa$ and die at background rate $\delta$, and residents presenting with COVID-19 symptoms (i.e. those in the Ich, Icl compartments) were assumed to be hospitalised at rate $\kappa_c$ and die at rate $\delta_c$ (see below for details).

Non-COVID-19 care home exit and entry processes were modelled stochastically as Poisson processes with identical rates. Therefore, without COVID-19, the care home was full at all times (100% occupancy). Non-COVID-19 care home exits (ie. background hospitalisation and mortality from compartments S, E, Ipc, Ia, R) were immediately replaced with entries to the care home from hospital. These entries included both residents returning from a hospital visit that was unrelated to COVID-19, and residents newly admitted to the care home from hospital. When COVID-19 symptomatic residents were present in the care home (Ich, Icl), their deaths were not replaced with new care home admissions from hospitals, as COVID-19 deaths occurred at a faster rate than replacement of residents, decreasing care home occupancy rates during the outbreak. Residents could die from COVID-19 both in the care home or in the hospital.

In the baseline scenario (medium community prevalence scenario), a mean of 5% of residents entering the care home from hospital were assumed to be recovered from SARS-CoV-2 infection all of whom entered the recovered (R) compartment, 0.07% residents entering the care home were assumed to be infected (of which 69% were exposed (E), 13% infectious pre-clinical (Ipc), 18% asymptomatic (Ia)). This was derived from combining the outputs of two mathematical models fit to English data from the community and to hospital data[15,30] (see Table S1 for details). The remaining residents entered the care home susceptible (S). We assumed that no residents who were hospitalised for reasons unrelated to COVID-19 entered the care home from hospital presenting with COVID-19 symptoms (Ich, Icl compartments).

Residents hospitalised for COVID-19 returned to the care home with a probability $p_{survival}$ (the probability that they survived their hospitalisation). In these simulations, therefore, the care home occupancy was further reduced due to COVID-19 deaths within the hospital. In the baseline scenario, on average 94% of residents who were admitted to hospital due to COVID-19 complications (Ich, Icl) and who survived their hospital stay were assumed to be fully recovered upon their return to the care home (direct re-entry to the R compartment). The remaining residents (on average 6%) were assumed to have a low level of infectiousness upon their return to the care home and were isolated upon re-entry. This assumption reflects that the tail of the viral load distribution beyond the average duration of hospital stays for COVID-19.[2–4]

Rates of hospitalisation and mortality have varied substantially during the pandemic.[31] Prior to the COVID-19 pandemic, nursing care home residents required but also received greater care, resulting in lower background rates of hospitalisation ($\kappa$) in nursing care homes than in residential care homes (0.84 vs. 1.13 per person per year in 2016/17[32]). Policy changes and changes in the perception of risk of hospitalisation have resulted in a decrease of the non-COVID hospitalisation rate for care home residents during the COVID-19 pandemic, particularly in those living in residential facilities.[31] In the baseline scenario (medium community prevalence), the rates of hospitalisation were 0.5 hospital visits per person per year for both types of facility, as extracted from SUS data (see below for details). The background mortality rate ($\delta$) was higher in nursing than in residential care homes (mean 0.4 vs. 0.2 per person per year, calculated using the number of non-COVID deaths in care homes published by the ONS[33], adjusted using estimates of lengths of stay prior to the COVID-19 pandemic from the literature, by type of facility[34], see below).

Not all care home residents with COVID-19 symptoms are hospitalised. The COVID-19 symptomatic hospitalisation rate ($\kappa_c$) was calculated by dividing the proportion of care home residents with COVID-19 who were hospitalised (mean 17%) by the duration of symptomatic infectiousness (mean five days) (for further details, see below). The hospitalisation rate for residents with COVID-19 clinical infection was assumed to be the same in residential and nursing care homes, as it was not evident that this rate would differ between settings. The COVID-19 symptomatic death rate $\delta_c$ was informed by an assumed mean case-fatality ratio of 25% (CFR literature estimates from care home residents range from 17%-36%[6–9,12,13]) for nursing care homes, and an assumed mean of 80% of these deaths occurring within the care home (20% of these deaths occurred during hospital visits, see below)[14].

##### *Hospitalisation rate for non-COVID-19 (κ)*

Weekly hospital admission rates from residential care homes and nursing homes from the week commencing 20 January to the week commencing 15 June 2020 were calculated using pseudonymised administrative data on hospital admissions from the Secondary Uses Service (SUS) database[35]. Care home residents were identified in SUS through linkage to address information from the National Health Applications and Infrastructure Services (NHAIS) database, which was matched to addresses of care homes registered with the Care Quality Commission (CQC), using validated linkage methodology. Elective and emergency hospital admissions to National Health Service (NHS) hospital trusts in England were included for individuals who had an address match to a care home in January 2020 and were still living in a care home at the time of admission. Hospital admissions were excluded if the administrative category was recorded as private patient, if the admission method was a transfer or was missing or if the patient classification for elective admissions was not recorded as day case or ordinary admission. To calculate hospital admission rates, admissions were also excluded if the primary diagnosis was suspected or confirmed COVID-19 (ICD-10 codes U07.1 and U07.2). Weekly admission rates per resident were calculated by across all residents for admissions from the week commencing 20 January to the week commencing 15 June 2020, controlling for the number of days spent in care homes across all residents. All processing of address information, and subsequent linkage of patient information, was carried out by the National Commissioning Data Repository (NCDR) and all data were anonymised in line with the Information Commissioner's Office (ICO)'s code of practice on anonymisation.

##### *Death rate from non-COVID-19 causes (δ)*

Care home resident deaths that were not due to COVID-19 (obtained by subtracting COVID-19 deaths from all-cause deaths) occurring within care homes, were extracted from the latest published ONS report, which reported deaths occurring until the 12 June 2020.[33] 7,378 non-COVID-19 deaths of care home residents occurring in care homes were reported for the period of the 14 May 2020 to the 12 June 2020 (the last 30 days of data available).

Assuming an occupancy of 80%, and that 11% of residents had COVID-19[25], the denominator population for the death rate was calculated as follows:
$N\, residents\, without\, COVID = (N\, beds\, registered \times occupancy) - (0.11 \times (N\, beds\, registered \times occupancy$
$= 460,000 * 0.8 - 0.11 * 460,000 * 0.8 = 327,520.$

The mean non-COVID-19 death rate, $\mu(\delta)$, was therefore given by:
$\mu(\delta) = 7,378/(30 * 327,520) = 0.00075$ per person per day, or 0.3 per year

Prior to the pandemic, the death rate in nursing care homes was higher than for residential care homes. Steventon et al. reported a length of stay in care homes of 283 for nursing care homes and 544.5 for residential care homes; and that 55% of nursing care home residents died within 30 days of leaving the facility, and 50% for residential care homes. Therefore, the mean pre-pandemic rates for nursing and residential care homes ($\mu(\delta_{pn})$and $\mu(\delta_{pr})$, respectively) were given by:
$\mu(\delta_{pn}) = (1/283) * 0.55 = 0.002$per person per day, or 0.7 per year
$\mu(\delta_{pr}) = (1/544.5) * 0.5 = 0.0009$per person per day, or 0.3 per year.
$\mu(m_{\delta rvn}) = \frac{\mu(\delta_{pr})}{\mu(\delta_{pn})}$
These mortality rates were similar to those reported by Shah et al. in 2013.[36]

The CQC register shows that approximately 50% of care homes were residential and 50% nursing.[37] Therefore, we adjusted the May/June death rates to give rates specific to nursing ($\delta_n$) and residential ($\delta_r$) care homes:
$\mu(\delta_n) = \frac{\mu(\delta)}{0.5\times\mu(m_{\delta rvn})+0.5} = \frac{0.00075}{0.5\times\frac{0.0009}{0.002}+0.5} 0.001$per day or 0.4 per year

$\mu(\delta_r) = \mu(\delta) \times \mu(m_{\delta rvn}) = 0.0005$per day or 0.2 per year

##### *COVID-19 symptomatic hospitalisation rate ($\kappa_c$)*

CFR literature estimates from care home residents range from 17%-36%[6–9,12,13]. For the purpose of this analysis we assumed a mean CFR of 25%. During the period of the 14th of May 2020 to the 12th of June 2020 (the last 30 days of data available, see above), approximately 80% of residents deaths occurred within the care home, and 20% in hospitals.[14] Therefore, we assumed that, on average, $0.25 \times 0.2 = 0.05$, 5% of COVID-19 residents died in hospital.

COVID-19 hospital mortality rates were calculated from SUS data for care home residents admitted between 18 May 2020 and 14 June 2020, using the destination on discharge variable. These data show a mean of 30% of patients admitted to hospital for COVID-19 died in hospital during the this period$(1 - p_{survival})$.

We calculated the mean probability of a resident with COVID-19 (symptomatic) being hospitalised from these data. We can illustrate the problem as follows:

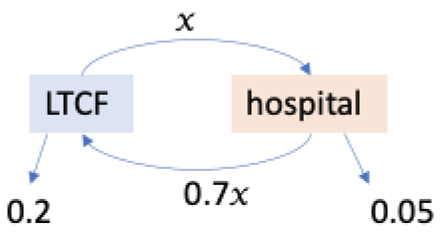


Where $x$ was the the mean probability of a resident with COVID-19 (symptomatic) being hospitalised and, on average, $p(resident\ with\ COVID - 19\ dying\ in\ hospital) = 0.05 = (1 - 0.7)x$

Therefore,
$$x = \frac{p(resident\ with\ COVID-19\ dying\ in\ hospital)}{1-p_{survival}} = \frac{0.05}{0.3} = 0.17$$

The COVID-19 symptomatic hospitalisation rate $\kappa_c$ was then calculated by dividing the proportion of care home residents with COVID-19 hospitalised (Beta distribution with a mean of 17%) by the duration of clinical infectioness (Gamma distribution with mean of 5 days).

##### *COVID-19 symptomatic death rate ($\delta_c$)*

The COVID-19 symptomatic death rate in nursing care homes ($\delta_{c,n}$) was calculated as follows:

$$\delta_{c,n} = \frac{CFR \times p_{death\ res\ within\ care\ home}}{duration\ \ of\ clinical\ infectiousness}$$

As most of the CFR studies were based in nursing care homes, this death rate was attributed to nursing care homes. The death rate for symptomatic residents in residential care homes ($\delta_{c,r}$) was calculated by adjusting this rate by the pre-pandemic ratio of the background mortality in nursing care home vs. residential care homes as follows:

$$\delta_{c,r} = \delta_{c,n} \times m_{\delta rvn}$$

#### SUPPLEMENTARY MATERIAL A3

Three community prevalence scenarios were considered: low, medium (baseline scenario), and high.

The community prevalence was derived from an established transmission model fit to community data[30]. We extracted the proportion of individuals in all ages that were susceptible (S), exposed (E), infectious preclinical (Ipc), infectious asymptomatic (Ia) and recovered (R) on the 2020-07-15 (low prevalence scenario), 2020-09-30 (medium prevalence scenario), and 2020-04-01. These were used to inform:

- the force of infection from visitors to residents ($\lambda_{v2r}$, through the probability that a visitor is infectious $p_{v_{infectious}}$, see Supplementary material A1),
- the force of infection from the community to staff ($\lambda_{c2s}$, through the probability of an individual in the community being infectious $p_{C_{infectious}}$),
- the proportion of replacement staff in each infectious state ($p_{S\,s2}$,$p_{EI\,s2}$, $p_{E,\,EI\,s2}$, $p_{Ipc,\,EI\,s2}$, $p_{Ia,\,EI\,s2}$, $p_{R\,s2}$), and
- the proportion of staff starting the simulation susceptible (S) and recovered (R).

We extracted the proportion of individuals aged over 70 that were susceptible (S), exposed (E), infectious preclinical (Ipc), infectious asymptomatic (Ia) and recovered (R) on the 2020-07-15 (low prevalence scenario), 2020-09-30 (medium prevalence scenario), and 2020-04-01. These were used to inform:

- The relative proportion of susceptible to recovered residents entering the care home from hospital after a hospitalisation unrelated to COVID-19.

The hospital discharge prevalence for those aged 65+ was derived from an established transmission model fit to hospital data[15]. We extracted the proportion of individuals that were susceptible (S), exposed (E), infectious preclinical (Ipc), infectious asymptomatic (Ia) and recovered (R) on the 2020-07-15 (low prevalence scenario), 2020-05-01 (medium prevalence scenario), and 2020-04-01. The early May hospital discharge data was used as a proxy for late September since the model output data was only available until mid-July (in early May the community prevalence was at a similar level as in late September). These data were used to inform:

- the proportion of residents entering each infectious state from hospital after a non-COVID-19 hospitalisation ($p_{S\,s2}$,$p_{EI\,s2}$, $p_{E,\,EI\,s2}$, $p_{Ipc,\,EI\,s2}$, $p_{Ia,\,EI\,s2}$, $p_{R\,s2}$).

The latest data point for the non-COVID-19 hospitalisation rate $\kappa$ available from SUS was for the week starting on 15 June (see Supplementary material A2 above). The adjusted emergency admissions for the overall population increased by 8% from June to July and by 11% from June to October (publicly available adjusted monthly NHSE data[38]). Assuming the same rate of increase in non-COVID-19-hospitalisation rates for care home residents, we adjusted the rate of the week starting the 15 June using the NHSE data to approximate the hospitalisation rate in mid-July (low community prevalence scenario) and late September (medium community prevalence scenario). The non-COVID-19 hospitalisation rate for residents in the high community prevalence scenario was extracted directly from SUS (see Supplementary material

A2 above) for the week starting on the 30 March. The late September approximation bore similar results to the SUS data for early May.

The reproduction number in the community $R_C$ was assumed to be 0.8 in the low community prevalence scenario, 1 in the medium community prevalence scenario, and 1.5 in the high community prevalence scenario. These were used to calculate the force of infection from the community to staff $\lambda_{c2s}$ (see Supplementary material A1 above for further details).

We also assumed that the probability of a care home experiencing an outbreak $p_{care\ home\ outbreak}$ would differ by community prevalence scenario. The $p_{care\ home\ outbreak}$ is used to calculate the force of infection to staff working in another care home $\lambda_{o2s}$. In the low community prevalence scenario we assumed 10% of care homes had an outbreak, in the medium community prevalence scenario we assumed 25% of care homes had an outbreak, and in the high community prevalence scenario we assumed 40% of care homes had an outbreak.

All other parameters were kept the same through the three community prevalence scenarios.

**SUPPLEMENTARY MATERIAL A4**

The impact of each strategy was assessed by estimating the cumulative probability of an outbreak (proportion of simulations with 1 or more infectious symptomatic residents in the care home), and of a large outbreak (proportion of simulations with 10 or more infectious symptomatic residents in the care home) over time. Eventually, every simulation resulted in an outbreak, therefore, we chose to compare the effectiveness of IPC interventions at two cut-offs: 30 days for the probability of an outbreak and 90 days for a large outbreak. Beyond 90 days it was unreasonable to assume the community prevalence scenario remained unchanged. The effectiveness of an intervention (i.e. the relative reduction in the probability of an outbreak) in preventing outbreaks at 30 days was calculated as follows:

$$Effectiveness\ of\ intervention\ in\ preventing\ an\ outbreak\ at\ 30\ days =$$

$$1 - \frac{cumulative\ probability\ of\ an\ outbreak\ at\ 30\ days\ under\ intervention}{cumulative\ probability\ of\ an\ outbreak\ at\ 30\ days\ under\ no\ intervention}$$

When assessing the effectiveness of testing, the comparison “no intervention” scenario referred to no testing, whilst it referred to the baseline assumptions when assessing the effectiveness of IPC interventions. Parameter sets were matched to enable the direct intervention comparison. The effectiveness of an intervention in preventing large outbreaks at 90 days was calculated in the same way. Approximately 3% of parameter sets yielded no large outbreaks at 90 days, and were discarded to enable the calculation of effectiveness.

#### REFERENCES APPENDIX


1 Lauer SA, Grantz KH, Bi Q, *et al.* The Incubation Period of Coronavirus Disease 2019 (COVID-19) From Publicly Reported Confirmed Cases: Estimation and Application. *Ann Intern Med* 2020; **172**: 577–82.
2 He X, Lau EHY, Wu P, *et al.* Temporal dynamics in viral shedding and transmissibility of COVID-19. *Nat Med* 2020: 1–4.
3 Ashcroft P, Huisman JS, Lehtinen S, *et al.* COVID-19 infectivity profile correction. *Swiss Med Wkly* 2020; **150**. DOI:10.4414/smw.2020.20336.
4 van Kampen JJA, van de Vijver DAMC, Fraaij PLA, *et al.* Shedding of infectious virus in hospitalized patients with coronavirus disease-2019 (COVID-19): duration and key determinants. *medRxiv* 2020. DOI:10.1101/2020.06.08.20125310.
5 Davies NG, Kucharski AJ, Eggo RM, Gimma A, Edmunds WJ, Centre for the Mathematical Modelling of Infectious Diseases COVID-19 working group. Effects of non-pharmaceutical interventions on COVID-19 cases, deaths, and demand for hospital services in the UK: a modelling study. *Lancet Public Health* 2020; **5**: E375–85.
6 Arons MM, Hatfield KM, Reddy SC, *et al.* Presymptomatic SARS-CoV-2 Infections and Transmission in a Skilled Nursing Facility. *N Engl J Med* 2020; **382**: 2081–90.
7 Graham NS, Junghans C, Downes R, *et al.* SARS-CoV-2 infection, clinical features and outcome of COVID-19 in United Kingdom nursing homes. *J Infect* 2020: 2020.05.19.20105460.
8 Patel MC, Chaisson LH, Borgetti S, *et al.* Asymptomatic SARS-CoV-2 infection and COVID-19 mortality during an outbreak investigation in a skilled nursing facility. *Clin Infect Dis* 2020; **71**. DOI:10.1093/cid/ciaa763.
9 Kennelly SP, Dyer AH, Martin R, *et al.* Asymptomatic carriage rates and case-fatality of SARS-CoV-2 infection in residents and staff in Irish nursing homes. *Age Ageing* 2021; **50**. DOI:10.1101/2020.06.11.20128199.
10 Dora AV, Winnett, Alexander, Jatt, Lauren P. Universal and Serial Laboratory Testing for SARS-CoV-2 at a Long-Term Care Skilled Nursing Facility for Veterans — Los Angeles, California, 2020. *MMWR Morb Mortal Wkly Rep* 2020; **69**. DOI:10.15585/mmwr.mm6921e1.
11 Kimball A. Asymptomatic and Presymptomatic SARS-CoV-2 Infections in Residents of a Long-Term Care Skilled Nursing Facility — King County, Washington, March 2020. *MMWR Morb Mortal Wkly Rep* 2020; **69**. DOI:10.15585/mmwr.mm6913e1.
12 McMichael TM, Currie DW, Clark S, *et al.* Epidemiology of Covid-19 in a Long-Term Care Facility in King County, Washington. *N Engl J Med* 2020: 1–7.
13 Dutey-Magni PF, Williams H, Jhass A, *et al.* Covid-19 infection and attributable mortality in UK Long Term Care Facilities: Cohort study using active surveillance and electronic records (March-June 2020). Infectious Diseases (except HIV/AIDS), 2020 DOI:10.1101/2020.07.14.20152629.
14 Number of deaths in care homes notified to the Care Quality Commission, England - Office for National Statistics. https://www.ons.gov.uk/peoplepopulationandcommunity/birthsdeathsandmarriages/deaths/datasets/numberofdeathsincarehomesnotifiedtothecarequalitycommissionengland (accessed June 25, 2020).
15 S Evans, Stimson J, Pople D, Bhattacharya A, White P, Robotham J. Quantifying the contribution of pathways of nosocomial acquisition of COVID-19 in English hospitals. *BMJ* (submitted).
16 Wikramaratna P, Paton RS, Ghafari M, Lourenco J. Estimating false-negative detection rate of SARS-CoV-2 by RT-PCR. *medRxiv* 2020: 2020.04.05.20053355-2020.04.05.20053355.
17 Chau NVV, Lam VT, Dung NT, *et al.* The natural history and transmission potential of asymptomatic SARS-CoV-2 infection. *medRxiv* 2020:

2020.04.27.20082347-2020.04.27.20082347.
18 Public Health England. Admission and care of residents in a care home during COVID-19. GOV.UK. https://www.gov.uk/government/publications/coronavirus-covid-19-admission-and-care-of-people-in-care-homes/coronavirus-covid-19-admission-and-care-of-people-in-care-homes (accessed Sept 28, 2020).
19 Public Health England. COVID-19: how to work safely in care homes. GOV.UK. https://www.gov.uk/government/publications/covid-19-how-to-work-safely-in-care-homes (accessed Sept 28, 2020).
20 Public Health England. Discharge into care homes: designated settings. GOV.UK. 2020. https://www.gov.uk/government/publications/designated-settings-for-people-discharged-to-a-care-home/discharge-into-care-homes-designated-settings (accessed Dec 22, 2020).
21 Department of Health and Social Care. Coronavirus (COVID-19): getting tested. GOV.UK. 2021. https://www.gov.uk/guidance/coronavirus-covid-19-getting-tested (accessed April 8, 2021).
22 Booth R. Government apologises for Covid testing delays at UK care homes. The Guardian. 2020; published online Sept 7. https://www.theguardian.com/world/2020/sep/07/uk-government-apologises-for-care-home-covid-testing-delays (accessed Sept 28, 2020).
23 Public Health England, NHS. Care home COVID-19 testing guidance - for testing staff and residents. 2021 https://assets.publishing.service.gov.uk/government/uploads/system/uploads/attachment_data/file/963633/Care_Home_Testing_Guidance_England_v22-02_2.pdf.
24 Department of Health and Social Care. Visiting care homes during COVID-19. GOV.UK. 2020; published online Dec 2. https://www.gov.uk/government/publications/visiting-care-homes-during-coronavirus/update-on-policies-for-visiting-arrangements-in-care-homes (accessed Dec 22, 2020).
25 Office for National Statistics. Impact of coronavirus in care homes in England. 2020. https://www.ons.gov.uk/peoplepopulationandcommunity/healthandsocialcare/conditionsanddiseases/articles/impactofcoronavirusincarehomesinenglandvivaldi/26mayto19june2020 (accessed April 8, 2021).
26 Pickering S, Batra R, Snell LB, *et al.* Comparative performance of SARS CoV-2 lateral flow antigen tests demonstrates their utility for high sensitivity detection of infectious virus in clinical specimens. *medRxiv* 2021: 2021.02.27.21252427.
27 Mahase E. Covid-19: Innova lateral flow test is not fit for “test and release” strategy, say experts. *BMJ* 2020; **371**: m4469.
28 Joint PHE Porton Down & University of Oxford SARS-CoV-2 LFD test development and validation cell. Preliminary report from the Joint PHE Porton Down & University of Oxford SARS-CoV-2 test development and validation cell: Rapid evaluation of Lateral Flow Viral Antigen detection devices (LFDs) for mass community testing. 2020 https://www.ox.ac.uk/sites/files/oxford/media_wysiwyg/UK%20evaluation_PHE%20Porton%20Down%20%20University%20of%20Oxford_final.pdf.
29 King AA, Nguyen D, Ionides EL. Statistical Inference for Partially Observed Markov Processes via the R Package pomp. *J Stat Softw* 2016; **69**: 1–43.
30 Davies NG, Kucharski AJ, Eggo RM, Gimma A, Group CC-19 W, Edmunds WJ. The effect of non-pharmaceutical interventions on COVID-19 cases, deaths and demand for hospital services in the UK: a modelling study. *medRxiv* 2020: 2020.04.01.20049908-2020.04.01.20049908.
31 The Health Foundation. Adult social care and COVID-19: Assessing the impact on social care users and staff in England so far. 2020 https://www.health.org.uk/publications/reports/adult-social-care-and-covid-19-assessing-the-i

mpact-on-social-care-users-and-staff-in-england-so-far (accessed April 8, 2021).
32 Wolters A, Santos F, Lloyd T, Lilburne C, Steventon A. Emergency admissions to hospital from care homes: how often and what for? The Health Foundation, 2019 https://www.health.org.uk/publications/reports/emergency-admissions-to-hospital-from-care-homes (accessed April 8, 2021).
33 Office for National Statistics. Deaths involving COVID-19 in the care sector, England and Wales. 2020; published online July 3. https://www.ons.gov.uk/peoplepopulationandcommunity/birthsdeathsandmarriages/deaths/datasets/deathsinvolvingcovid19inthecaresectorenglandandwales (accessed July 7, 2020).
34 Steventon A, Roberts A. Estimating length of stay in publicly-funded residential and nursing care homes: A retrospective analysis using linked administrative data sets. *BMC Health Serv Res* 2012; **12**: 1–1.
35 Grimm F, Hodgson K, Brine R, Deeny SR. Hospital Admissions From Care Homes in England During the COVID-19 Pandemic: A Retrospective, Cross-Sectional Analysis Using Linked Administrative Data. *Preprints* 2021. DOI:10.20944/preprints202102.0593.v1.
36 Shah SM, Carey IM, Harris T, DeWilde S, Cook DG. Mortality in older care home residents in England and Wales. *Age Ageing* 2013; **42**: 209–15.
37 Care Quality Commission. Active locations for providers registered under the Health and Social Care Act (HSCA). 2020. https://www.cqc.org.uk/about-us/transparency/using-cqc-data (accessed April 8, 2021).
38 NHS England. A&E Attendances & Emergency Admission statistics, NHS and independent sector organisations in England. 2020 https://www.england.nhs.uk/statistics/statistical-work-areas/ae-waiting-times-and-activity/ae-attendances-and-emergency-admissions-2020-21/.
